# Radiologist observations of chest X-rays (CXR) predict sputum smear microscopy status in TB Portals, a real-world database of tuberculosis (TB) cases

**DOI:** 10.1101/2022.04.21.22273975

**Authors:** Gabriel Rosenfeld, Andrei Gabrielian, Alyssa Meyer, Alex Rosenthal

## Abstract

The Tuberculosis (TB) Portals is an international program of 14 countries connecting clinical, genomic, and radiologist specialists to develop an openly available repository of deidentified TB cases with multi-modal data such as case clinical characteristics, pathogen genomics, and radiomics. This real-world data resource contains over 4000 TB cases, principally drug resistant cases, with over 4000 chest X-rays (CXR) images. The scope of curated data offers a case-focused perspective into the drivers of disease incorporating the chronological context of the presented CXR data. Here, we analyze a cohort consisting of new TB cases to understand the relationship between baseline sputum microscopy status and nearby Chest X rays images. The Timika score, a lung biomarker of disease severity, was derived for each CXR using available radiologist observations. The Timika score along with the radiologist observations were compared for predictive performance of baseline sputum microscopy status. Baseline sputum microscopy status is a useful marker of pre-treatment disease severity and infectiousness. The modeling results support that both the radiologist observations as well as Timika score are predictive of smear status and that Timika score performs similarly to the top 5 radiologist features by feature selection. Moreover, inferential statistical analysis identifies the factors having the greatest association with sputum smear positivity such as presence of radiologist observations in both lungs, presence of cavity, presence of nodule, and Timika score itself. The results are consistent with prior reports showing Timika Score utility for predicting baseline sputum smear and disease status. We report testing of Timika Score on the largest, openly available real-world dataset of TB cases that can serve as a reference to explore extant and new TB disease severity scores bridging radiological, microbiological, and clinical data. To illustrate, we visualize Timika score from images in our database with other cases characteristics demonstrating that this score captures lung biomarker status consistent with known clinical risk factors.

## Introduction

Tuberculosis (TB) remains a major global pandemic with approximately 10 million new cases and 1.5 million deaths each year (1, 2). With the emergence of the SARS-Cov2 global pandemic in 2020, it is estimated that the TB pandemic may have worsened due to additional strains and challenges encountered via healthcare systems around the world (3, 4). Concurrent to those unfortunate events, drug resistant TB continues to be a persistent threat with up to ∼20% of TB isolates around the world estimated to be resistant to a major drug. Transmission of drug resistant TB is an emerging phenomenon closely monitored by health authorities worldwide (5). Drug resistant TB cases (DR-TB) are associated with poorer outcomes and more expensive cost of care when compared to drug sensitive TB. DR-TB has a lower treatment success of approximately 55% globally and Multi- or Extensively DR-TB care can cost up to 25 times that of TB cases that are drug sensitive (6, 7). Therefore, real-world databases focusing on these DR cases that span multi-domain case information are essential to identify novel relationships and aspects of drug resistance to enable translational medicine to timely and efficiently address drug resistance.

To eradicate TB, clinicians need rapid diagnostics of disease along with efficient means of monitoring treatment response and completeness at discharge. Sputum smear microscopy has been a primary method for diagnosis of pulmonary tuberculosis in low and middle income countries (LMIC) since it is a relatively simple, rapid, and less costly approach that can identify the most infectious patients and be applied in a variety of socio-economic status areas. Nonetheless, this approach shows deficiencies in certain demographic groups such as extra-pulmonary TB, pediatric TB, and TB patients simultaneously infected with HIV (8). Moreover, the requirement for repeated sputum sampling can present obstacles to the application of the approach as patients may not return for results, are lost to follow up, or have difficulty producing usable sputum samples. Despite these challenges, it is still widely used throughout LMIC for disease monitoring and response to therapy (9) as well as having demonstrated some ability to predict treatment response albeit requiring additional clinical factors (10). Since this microbiological information is sometimes unavailable or inclusive, it is important to identify other modalities that may assist with diagnosis or monitoring of treatment response, and one such approach is imaging of the lungs via Chest X-rays (CXRs).

CXR imaging is often collected during TB disease management to understand treatment response and disease status. Unlike computed tomography (CT) imaging that may not always be available due to the cost of associated infrastructure (11), CXRs are the primary means of assessing lung status in LMIC due to their relatively lower costs (12). As such, they are more widely available to clinicians for assessing lung status during TB disease management and used as a decision-making clinical information point compared to CTs. Radiologist assessment of CXRs have been the gold standard reference upon which CXRs have been interpreted for clinical decision making historically. These observations provide an important lung biomarker that can inform patient risk, disease severity, and response to treatment over the course of a TB case. For example, Heo et al. tracked radiological lesions from CXRs over the course of TB treatment in a prospective cohort analysis showing that presence of cavity or fibrotic lesion associated with poor radiological response (13). Another example is the development of CXR-derived Timika Score that has been associated with baseline sputum smear microscopy status and disease severity in TB cases (14, 15).

The National Institute of Allergy and Infectious Diseases (NIAID) Office of Cyber Infrastructure and Computational Biology leads the transnational partnership of participating sites covering 14 countries with heavy DR TB burden. This partnership created the TB Portals to facilitate TB data sharing and science with a goal towards a better understanding of the real-world aspects of especially problematic DR TB. The TB Portals resource consists of a repository of TB case data including multiple domains such as case clinical characteristics, pathogen genomics, and radiomics that can support the biomedical research community’s research efforts towards TB. As of April 2021, the TB Portals database includes over 4000 TB cases, mostly drug resistant cases, with over 4000 CXRs. Many of these cases also have radiologist annotations for their CXRs to assess lung biomarker status in relation to the clinical and microbiological characteristics of the case. While other resources have large numbers of chest X-ray images, TB Portals provides a TB case-centered repository encapsulating the chronological context associated with the CXR such as drug resistance status, regimens administered so far, the genome of the pathogen, and sputum microscopy status. External collaborators can apply for access to publicly shared data through an online data use agreement (DUA) and then download this data to facilitate reproducibility and open science.

In this study, we utilize the radiologist observations for CXR images in the TB Portals repository to derive Timika Score (15), a useful numerical lung biomarker, to assess its utility for predicting sputum smear microscopy status. We compare Timika Score performance with other features we derived from the radiologist-reported observations to determine if the additional features could improve upon Timika’s previously reported performance. We select a cohort of cases with a case definition of new containing sputum smear microscopy results from specimens taken prior to start of treatment, as well as CXRs with radiologist observations within two weeks of the specimen date. We perform inferential statistical analysis of risk of positive sputum microscopy from presence of various features derived from radiologist observations. We also examine Timika Score in relation to other aspects of the case such as demographics, case definition, and outcome. For instance, we utilize a strength of this resource in having a larger number of mono drug resistant (Mono DR), poly drug resistant (Poly DR), Multi-drug resistant (MDR) and Extensively drug resistant (XDR) according to WHO guidelines (16).

We report results consistent with prior publications regarding the utility of the Timika score for predicting baseline sputum status. Importantly, we show that Timika Score offers similar predictive performance compared to the top 5 features we derive from radiologist observations. These results suggest that Timika Score is well-optimized for determining pre-treatment disease infectiousness and severity status.

## Materials and Methods

### Computing environment

All analyses were done on a MacBook Pro laptop (x86_64-apple-darwin15.6.0 (64-bit) Running under: macOS Mojave 10.14.6) using R version 4.0.2 (2020-06-22) and RStudio 1.2.5033. Specific versions of the R packages can be found in the associated code which contains renv.lock file listing all used packages and version numbers.

### Cohort selection

#### Sputum Prediction

To remove the potential of confounding lung biomarkers due to prior history of TB, new cases of TB with CXRs containing radiologist annotations as well as sputum microscopy test results from specimens prior to or on the treatment start date were selected. Those cases where the specimen collection date was within 14 days of a CXR were included. For cases where multiple pairs of specimens and CXRs existed, the last specimen prior to treatment was used. For cases where multiple microscopy test results were present for a specimen, the last microscopy test result for that specimen was used. For cases where multiple images existed, the last imaging date was used. Unknown or non-standard microscopy results such as “Unknown data” and “Saliva” were excluded. Code used for generating the cohort is provided in the Data availability and code section. Ultimately, 572 new cases were selected from the database with sputum microscopy results of Negative, 1 to 9 in 100 (1-9/100), 10 to 99 in 100 (1+), 1 to 9 in 1 (2+), 10 to 99 in 1 (3+), and More than 99 in 1 (4+) consisting of 259, 29, 144, 60, 58, and 22 cases respectively within this cohort. The cohort characteristics summarizing case details for the sputum prediction cohort can be found in Supplementary Table 1.

#### Analysis of Timika Score with regards to other case characteristics in TB portals

For Figure 1 and Figure 2 visualizing the Timika Score in relation to other case characteristics, we used all available images with manual radiologist annotations from the February 2021 release of TB Portals data available for download from Aspera. This included 2058 images from 1761 cases covering not just New cases but all other types in the database. The characteristics summarizing corresponding case details for the set of available images for the Timika Score visualizations can be found in Supplementary Table 2.

**Fig 1.**
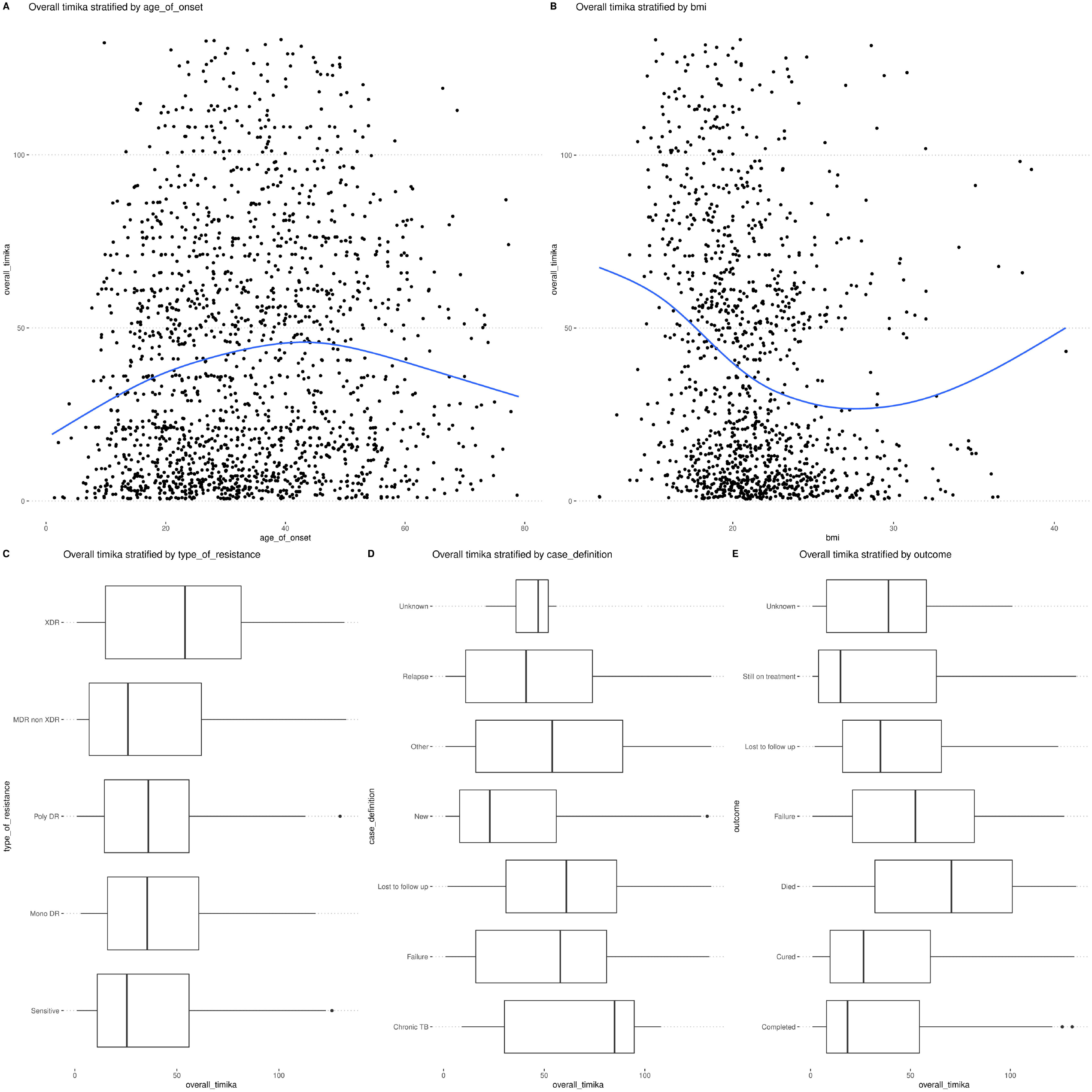
Association of Timika Score from initial CXR with radiologist observations with other case attributes. Timika Score derived from initial available CXRs associated with cases in the TB Portals repository are visualized along with a variety of salient case characteristics with missing observations dropped according to variable (N = 1757). For A) and B), the age of onset (N = 1757) and BMI (N = 1268) from the case are shown with blue trend line respectively. For C), D), and E), boxplots with interquartile range showing Timika Score by the type of drug resistance, status of case at start, and status of case at end are shown. In C), MDR non XDR (N = 752), Mono DR (N = 118), Poly DR (N = 78), Sensitive (N = 514), and XDR (N = 295) case drug resistance statuses are shown with the associated Timika Score from initial CXR with the case. XDR cases tend to show relatively higher Timika Scores. In D), Chronic TB (N = 18), Failure (N = 179), Lost to follow up (N = 65), New (N = 1141), Other (N = 45), Relapse (N = 304), and Unknown (N = 5) case definitions are shown with the associated Timika Score from initial CXR with the case. Undesirable case definitions such as Failure, Lost to follow up, Relapse, Chronic TB, or Unknown from prior history show higher Timika Score compared to New cases. In E), Completed (N = 170), Cured (N = 984), Died (N = 126), Failure (N = 128), Lost to follow up (N = 151), Still on treatment (N = 169), and Unknown (N = 29) case outcomes are shown with the associated Timika Score from initial CXR with the case. Undesirable outcomes such as Died, Failure, Lost to follow up, or Unknown show higher Timika Score compared to beneficial outcomes such as Completed, Cured, or Still on treatment.

**Fig 2.**
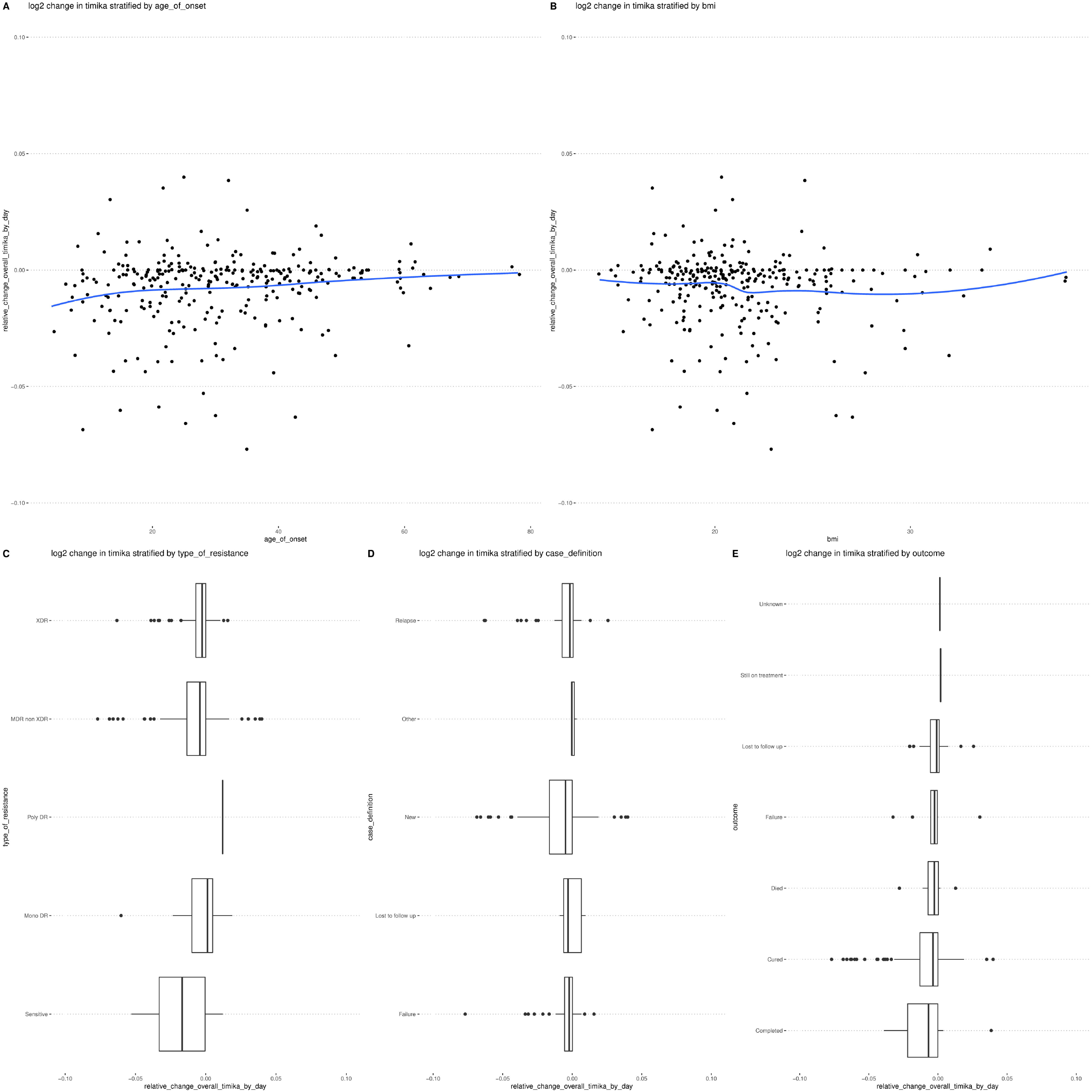
Association of relative change in Timika Score by day from initial CXR with radiologist observations to last available CXR with radiologist observations with other case attributes. Log2 relative change in Timika Score by day from initial available CXR to last available CXR associated with cases in the TB Portals repository are visualized along with a variety of salient case characteristics with missing observations dropped according to variable (N = 297). For A) and B), the age of onset (N = 297) and BMI (N = 292) from the case are visualized with log2 relative change in Timika Score by day with blue trend line respectively. To aid in visualizing the trendline, the y axis was limited to between −0.1 and 0.1 resulting in an additional 9 outliers being removed for age of onset and BMI case numbers above respectively. For C), D), and E), boxplots with interquartile range showing log2 relative change in Timika Score by day compared by the type of drug resistance, status of case at start, and status of case at end are shown. In C), MDR non XDR (N = 160), Mono DR (N = 13), Poly DR (N = 1), Sensitive (N = 24), and XDR (N = 99) case drug resistance statuses are shown with the log2 relative change in Timika Score by day from initial CXR to last available CXR. To aid in visualization, y axis was limited to −0.1, and 0.1 resulting in additional 5, 2, 0, 2, and 0 outliers being removed from MDR non XDR, Mono DR, Poly DR, Sensitive, and XDR case numbers above respectively. In general, drug resistant cases show lower relative change in Timika Score although several groups show low N and must be interpreted cautiously. In D), Failure (N = 35), Lost to follow up (N = 8), New (N = 184), Other (N = 4), and Relapse (N = 66) case definitions are shown with the log2 relative change in Timika Score by day from initial CXR to last available CXR. To aid in visualization, y axis was limited to −0.1, and 0.1 resulting in additional 0, 0, 8, 1, and 0 outliers being removed from Failure, Lost to follow up, New, Other, and Relapse case numbers above respectively. Deleterious case definitions such as Failure, Lost to follow up, Relapse, or Other from prior history show less change in relative Timika Score compared to New cases. In E), Completed (N = 27), Cured (N = 223), Died (N = 15), Failure (N = 10), Lost to follow up (N = 20), Still on treatment (N = 1), and Unknown (N = 1) case outcomes are shown with the log2 relative change in Timika Score from initial CXR to last available CXR. To aid in visualization, y axis was limited to −0.1, and 0.1 resulting in additional 4, 5, 0, 0, 0 and 0 outliers being removed from Completed, Cured, Died, Failure, Lost to follow up, Still on treatment, and Unknown case numbers above respectively. Deleterious outcomes such as Died, Failure, Lost to follow up, or Unknown show smaller relative changes compared to beneficial outcomes such as Completed and Cured. As above for other visualizations, caution is warranted given the small N’s associated with certain subgroups.

### Data preprocessing and extraction of feature set

Data from TB portals was downloaded as a list of .csv files from the Aspera file share service using the February 2021 version of each respective file. The CXR manual annotations are provided as a set of features corresponding to observations by radiologists within each sextant of the lungs (dividing each lung by 1/3) as well as a set of features provided at the level of the entire lung. For those features corresponding to sextant level observations, features where no observations were provided by radiologists were imputed as 0 (for numerical data corresponding to between 0-100% involvement of sextant) or “No” (for categorical data indicating presence/absence of a specific feature within the sextant). The omission of these at either the level of the entire sextant or specific sextant-level feature are interpreted as the radiologist did not observe the feature.

After imputation, features were converted for tidy-like data processing using packages from the R tidyverse. This permits various types of downstream feature engineering such as identification of involvement of one or both lungs by sextant-level feature type, calculation of summary statistics for numerical features across sextants, and other score calculations such as Timika Score. For this analysis, involvement of both lungs as well as mean percentage of sextant involvement across all sextants by specific sextant-level radiologist observation was calculated along with Timika Score. Both lung features were calculated in a binary manner where involvement of a left and right sextant for the features was required to indicate involvement of both lungs for that feature. Timika Score was calculated like the original publication (15) by a simple method of taking the overall abnormal percent of volume of the lungs reported by the radiologist and adding 40 if the presence of cavity was indicated in the radiologist report. Characteristics of derived features by microscopy status can be found in Supplementary Table 3.

MLR3 framework was used to define a set of prediction tasks as well as pipelines for modeling (17). 70% of the data was selected as a training set and 30% was held out as a validation set. We tested two distinct prediction tasks in the MLR3 framework: 1) to predict sputum positive (1 to 9 in 100, 1+, or higher) compared to negative and 2) to predict higher bacterial load positive (2+, 3+, or higher) compared to negative. The positive to negative prediction task was relatively well balanced between classes; however, the high bacterial load positive to negative prediction task showed moderate class imbalancing so a class balancing step was included in some pipelines for comparison. Machine learning (ML) pipelines using all derived radiological features were compared to pipelines using only the Timika Score for prediction. For ML pipelines using all derived radiological features, factor data was encoded to a binary indicator (https://mlr3pipelines.mlr-org.com/reference/mlr_pipeops_encode.html), low variance features were removed (https://mlr3pipelines.mlr-org.com/reference/mlr_pipeops_removeconstants.html), features were scaled by min-max scaling (https://mlr3pipelines.mlr-org.com/reference/mlr_pipeops_scalerange.html), and the top 5 features were selected via a variety of feature selection methods (https://mlr3filters.mlr-org.com/) or Principal Component Analysis (https://mlr3pipelines.mlr-org.com/reference/mlr_pipeops_pca.html). The following ML models were assessed as part of the pipelines: featureless, glmnet, kknn, multinom, naïve bayes, rpart, ranger, xgboost, svm, and nnet as described in the subsequent link (https://mlr3learners.mlr-org.com/reference/index.html). For pipelines using only Timika score, only the min-max scaling step was included as part of the pipeline.

### Model performance benchmarking

5-fold cross validation was used to assess various binary classification metrics towards respective prediction tasks on the training set. Both the metrics and the resampling strategies can be found in the mlr3 documentation (https://mlr3.mlr-org.com/reference/index.html) under Measures and Resampling Strategies sections. We also assess these binary classification metrics in the validation dataset to ascertain performance on data which has never been used during model training. To compare performance of the top radiologist observations and Timika Score, the best pipelines using the top radiologist derived features were compared by bootstrapping without replacement (N = 200) to the best pipelines using Timika Score alone, or a featureless pipeline as a control to indicate the density of observed model performance particular to this dataset that may be due to random chance.

### Calculation of inferential statistics

To estimate the univariate Odds Ratios (OR) and multivariate adjusted Odds Ratios (AOR), the finalfit R package was used. Both_hugenodule1 feature was removed from the analysis as it did not show any variance. As Timika Score is highly correlated with other variables in the dataset (e.g. overall abnormal volume and cavity), MRMR feature selection (https://mlr3filters.mlr-org.com/reference/mlr_filters_mrmr.html) was performed for the top 5 features to include in the multivariate modeling. As both_hugenodule1 feature was excluded, less than 5 features were selected in the multivariate modeling. The both_lungs feature was included in the multivariate model to adjust for indication of involvement of both lungs from sextant level features when assessing estimated odds of sputum positivity.

### Visualization of Timika Score with other case attributes

Interesting case variables were examined for association with Timika Score to assess consistency with current understanding of TB clinical risk factors. This included demographic information such as age and BMI as well as case resistance status, case definition, and treatment outcome. The initial CXR with available radiologist observations were selected for each case, Timika Score calculated and plotted using ggplot2 to visualize associations with other case attributes.

For calculating temporal changes in Timika Score, those cases with an initial CXR identified above were filtered for cases with an additional follow up CXR with radiologist observation. The log2 transformed relative change were calculated for each image’s Timika Score comparing the earliest score with all subsequent scores. To account for differences in the length of time between images that may impact the relative score change, the difference in number of days between CXRs was calculated and the log2 transformed relative changes were divided by the number of days between pairs of images for each case to generate the log2 relative change by day.

### Data availability and code

The TB portals program necessitates all users of the data sign a DUA before access to the underlying, de-identified clinical data is provided and the data can be requested at the following URL (https://tbportals.niaid.nih.gov/download-data). Therefore, this study provides the code to reproduce the analysis without the underlying raw data (https://github.com/niaid/tbportals.xray.sputum.2021) in compliance with the DUA. To rerun the analysis, interested parties can request data access by completing the DUA and then place the downloaded files to the subdirectory of the data folder as provided in the GitHub repo instructions. To aid reproducibility, the list of patient IDs and condition IDs used from the sputum prediction analysis are provided in Supplementary Table 4. The specific record identifiers for the set of images used for visualization of Timika Score in comparison with other case attributes are provided in Supplementary Table 5. For cases where change in Timika was calculated over time, the first and last images used in the case are shown along with the dates ofthe image. Both supplementary tables allow those interested in examining the specific records to do so after completion of required DUA irrespective of the evolution in number of available cases in the database.

## Results

### Timika score associates with case clinical characteristics, disease severity, and risk

The TB portals resource contains case information bridging across domains of interest such as clinical, demographic, radiologic, and pathogen genomics. We leveraged the unique value of these connections to assess any scores or other features of interest from the derived radiological data. After generating the Timika Scores from all available CXRs with associated radiologist annotations, we explored the relationship between Timika Score with the additional information contained about the case mentioned above. We analyzed these relationships to determine if they are consistent with prior TB clinical findings to assess the plausibility of the derived radiological data.

#### Age, BMI, Type of Resistance, Case Definition, and Case Outcome show associations with Timika Scores

We used only the initial image with associated radiologist observations for the visualizations assessing Timika Score in relation to other aspects of the case. We visualize Timika Score with age of onset, BMI, resistance type, case definition, and case outcome and include a trendline in the relationships for any numerical features. We identify relationships between Timika score and case characteristics of interest that are consistent with our prior knowledge of TB clinical risk.

For instance, Timika Score of the initial image gradually increases with age of onset until the relationship plateaus around age 50 and decreases although some of the decrease and initial increase can likely be attributed to the lower density of observations at the two extremes of age (Fig 1A). Timika Score decreases with increasing BMI until it plateaus around a BMI of ∼25 (Fig 1B). Like age, the extremes of the BMI visualization need to be interpreted cautiously given the lower densities. XDR cases, resistant to the most TB drugs and with the worst outcomes, are observed to have a higher median Timika Score and interquartile range compared to other case resistance types (Fig 1C). New cases of TB are shown to have a lower median Timika Score and interquartile range compared to other types of cases such as Chronic TB, Prior treatment failure, Relapse, Prior lost to follow up, or Other prior unknown status case definitions (Fig 1D). Similarly, visualizing case outcomes reveals that detrimental treatment outcomes such as Died, Treatment failure, Lost to follow up, or Unknown show higher median Timika Scores compared to beneficial outcomes such as Treatment completion, Cured, or Still on treatment (Fig 1E). Taken together, the results demonstrate associations between Timika Score from initial available image and case characteristics that reflects TB clinical risk.

#### Age, BMI, Type of Resistance, Case Definition, and Case Outcome show associations with the temporal changes in Timika Scores

Of those cases with initial CXR images visualized above, we next examined changes in Timika Score whenever follow up CXR images were available. To do so, we filter on cases with this additional imaging information. We calculate log2 transformed Timika Score from initial image to last available image per case dividing by the number of days between images to account for the relative amount of time between each image. We use ggplot2 to visualize log2 transformed change in Timika Score by day with age of onset, BMI, resistance type, case definition, and case outcome and include a trend line in the associations for any numerical features. Interestingly, most cases have a negative relative change in Timika Score by day indicating improvement in lung status over the course of the case. Such a decrease over time would be expected given these cases would have been undergoing clinical management. We observed associations between the relative change by day and case characteristics of interest that are consistent with prior knowledge of TB clinical risk.

For instance, the log2 transformed change in Timika Score by day steadily decreases with age of onset such that younger age shows greater relative change whereas older age shows less relative change (Fig 2A). Conversely, log2 transformed change in Timika Score by day increases as BMI increases (Fig 2B). Lower BMI demonstrates a smaller relative change as compared to higher BMI. When examining relative change by resistance type of the case, Drug sensitive cases are observed to have a larger relative change compared to drug resistant types (Fig 2C). Similarly, new cases of TB show greater relative change compared to other types of cases such as Prior treatment failure, Prior lost to follow up, Relapse, or Other prior status at the start of the case (Fig 2D). Visualizing by case outcomes demonstrates that undesirable outcomes such as Died, Treatment failure, or Lost to follow up show decreased relative change by day compared to beneficial outcomes such as Treatment completion or Cured (Fig 2E). Like earlier visualizations of the Timika Score from available initial image, observations of relative changes are consistent our prior understanding of TB clinical risk factors.

#### Sputum smear microscopy results associate with Timika score in this cohort of TB portals cases

We next assessed the previously reported role of Timika Score for predicting sputum smear status by analyzing the selected cohort of new cases having microscopy results from sputum specimens taken prior to treatment with associated images within two weeks of the specimen (N = 572). Mean Timika Score is lowest amongst new cases with a sputum microscopy result of negative and increases for microscopy results indicating a higher burden of bacteria within sputum [1 to 9 in 100, 1+, 2+, etc.] (Fig 3). Only the 4+ level shows slightly lower mean Timika Score compared to the next highest level of 3+, which may be due to variance from the lower number of available cases in this 4+ level (N = 22). This clear trend in the TB portals dataset is consistent with previously reported role of Timika Score for predicting baseline sputum status.

**Fig 3.**
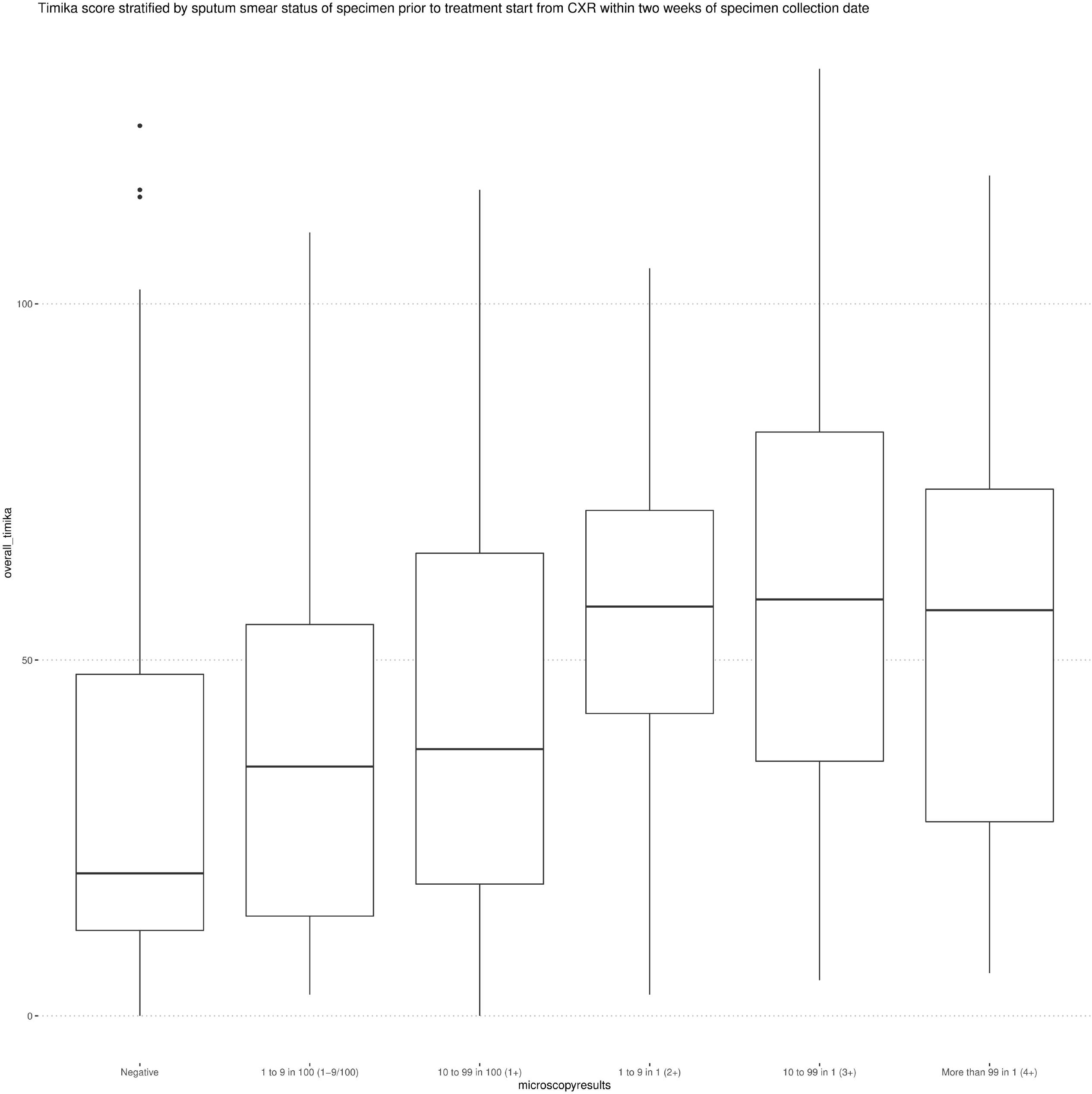
Timika score derived from radiologist observations of CXR within two weeks of specimen taken prior to treatment start. Timika score is visualized by the smear microscopy results of specimens taken prior to treatment start for which CXRs were available within two weeks of specimen date (N = 572). Boxplots show median Timika Score for associated images with interquartile range. Images associated with negative smear microscopy status have lower Timika Scores while those with positive statuses (1 to 9 in 100, 1+, and higher) show progressively higher Timika Scores that appear to plateau around 2+ or higher. The following number of Timika Scores derived from radiologist observations of images are available for Negative, 1 to 9 in 100 (1−9/100), 10 to 99 in 100 (1+), 1 to 9 in 1 (2+), 10 to 99 in 1 (3+), and More than 99 in 1 (4+) groups respectively: 259, 29, 144, 60, 58, 22.

### Inferential statistics associated with sputum microscopy status

Given the association of Timika Score with sputum smear microscopy, we continued our investigation by assessing the risk of a positive sputum microscopy status (1 to 9 in 100, 1+, or higher) compared to a negative status using Timika score along with the other derived features from radiologist observations. To do so, we performed univariate and multivariate logistic regression removing any feature with no variance that caused univariate or multivariate modeling to fail. We leveraged MRMR feature selection to select the top 5 features for multivariate models. Timika Score is derived from the radiological features (e.g., presence of cavity and overall abnormal volume) so we wanted to select additional features that would not directly correlate with Timika Score but still potentially correlate with sputum microscopy status.

We observe multiple features with evidence of involvement of both lungs showing higher risk of sputum microscopy positivity including calcified nodules, fibrotic nodules, low density nodules, involvement of both lungs by indication of any type of sextant feature, medium density nodules, medium cavities, small cavities, multiple cavities, and small nodules (Table 1). For numeric variables, large cavities, low density nodules, medium cavities, small cavities, and overall percent of abnormal volume showed statistically significant increases in risk of positive sputum result (active pathogen detected in the sputum) per each unit increase in percentage whereas pleural effusion percent of hemithorax involved showed the opposite. Each unit increase in Timika Score showed an increased risk in pre-treatment sputum positive microscopy status consistent with its prior reported role. In the multivariate model, the Timika Score showed a higher risk of positive sputum microscopy status after adjusting for indication of involvement of both lungs. This suggests that risk of positive sputum microscopy status does not require evidence of both lung involvement but rather greater percentage abnormal regions or cavity area may be sufficient for the increased risk indicated by Timika Score. Interestingly, the other MRMR selected features of the multivariate model were all indicators of involvement of both lungs for the respective features. Only the indication of calcified nodules in both lungs demonstrated a statistically significant increase in risk of positive sputum microscopy status adjusting for other covariates in the multivariate model. This feature could suggest cases with unreported prior history of pulmonary TB.

**Table 1.**
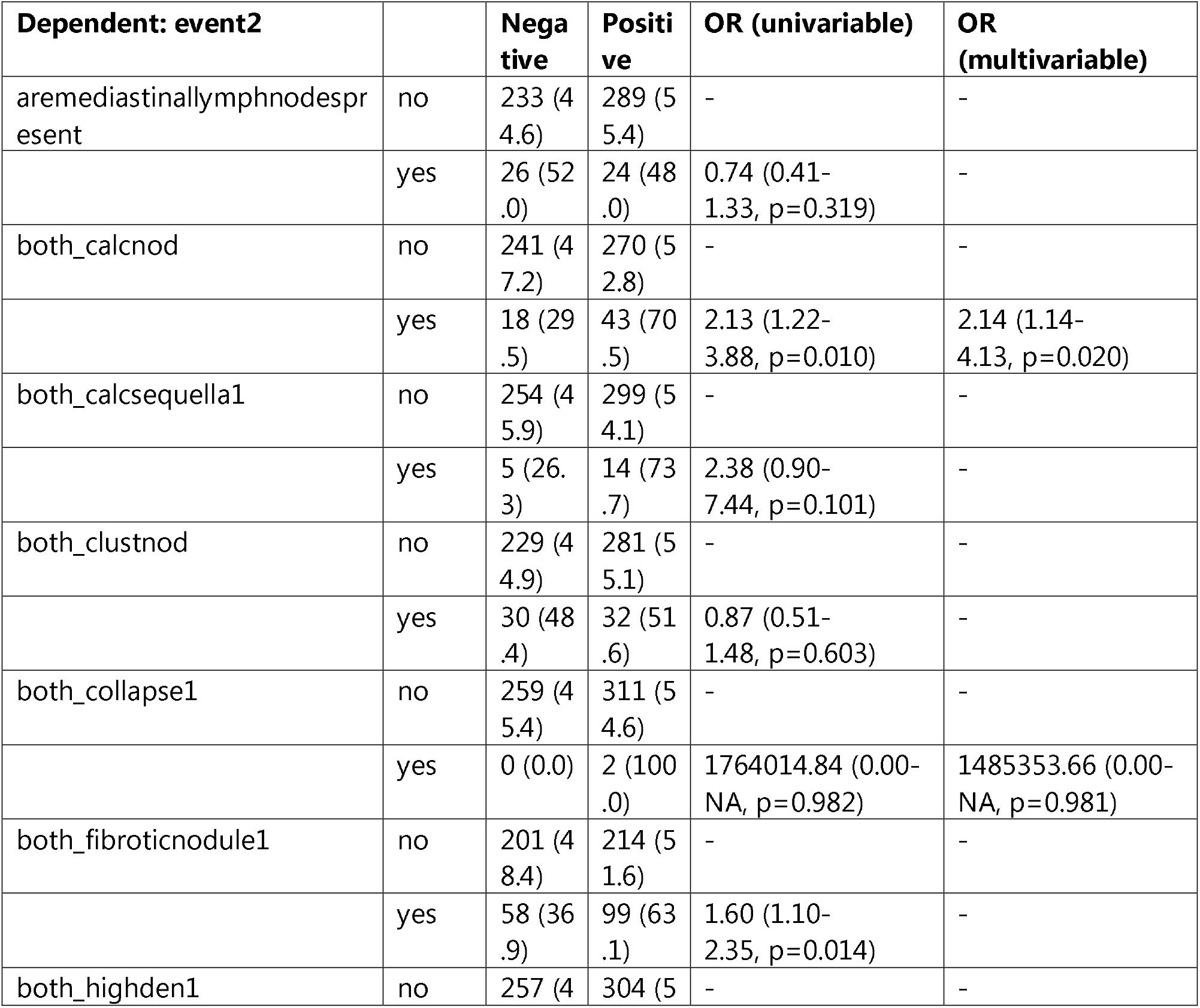
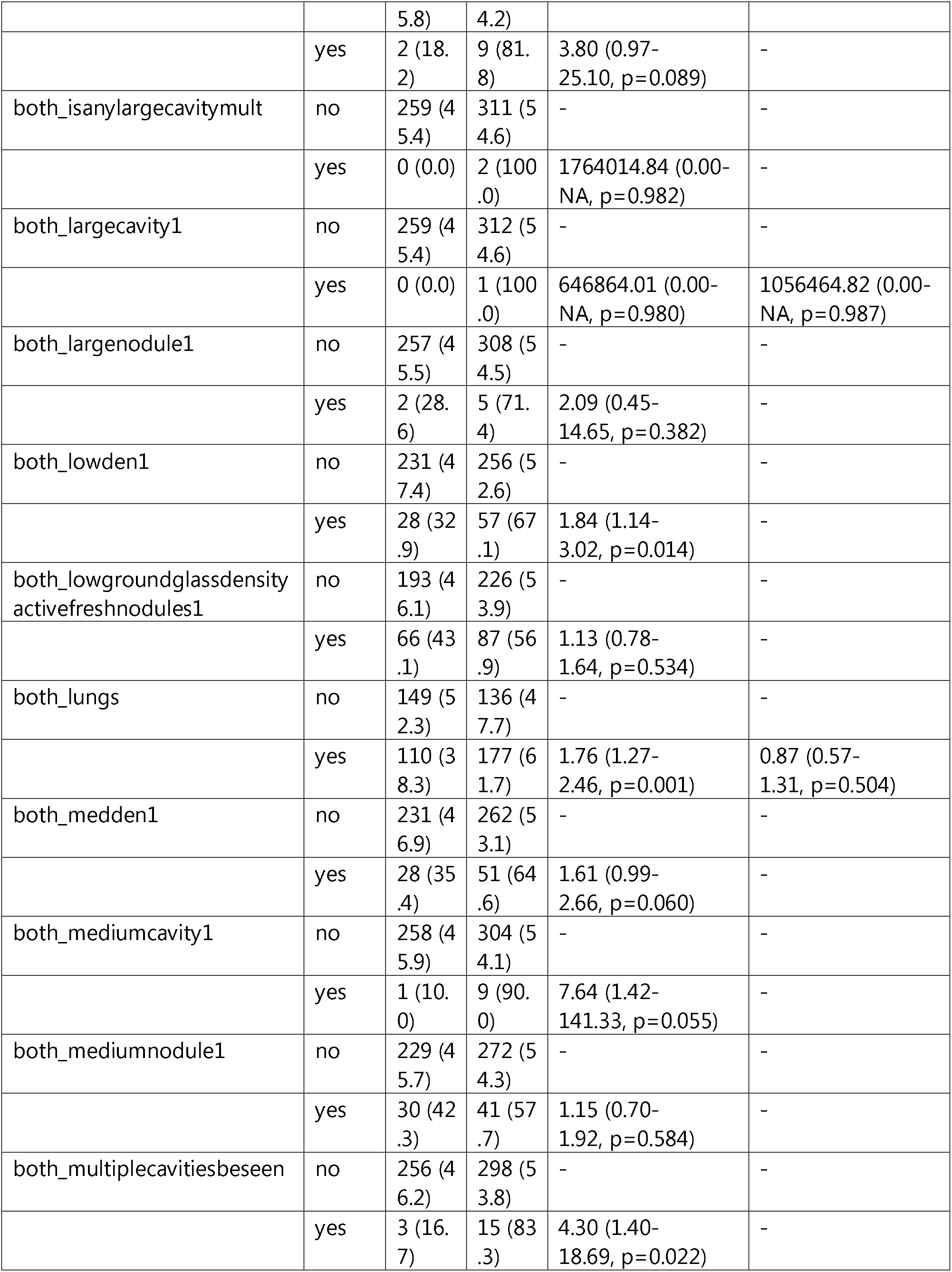
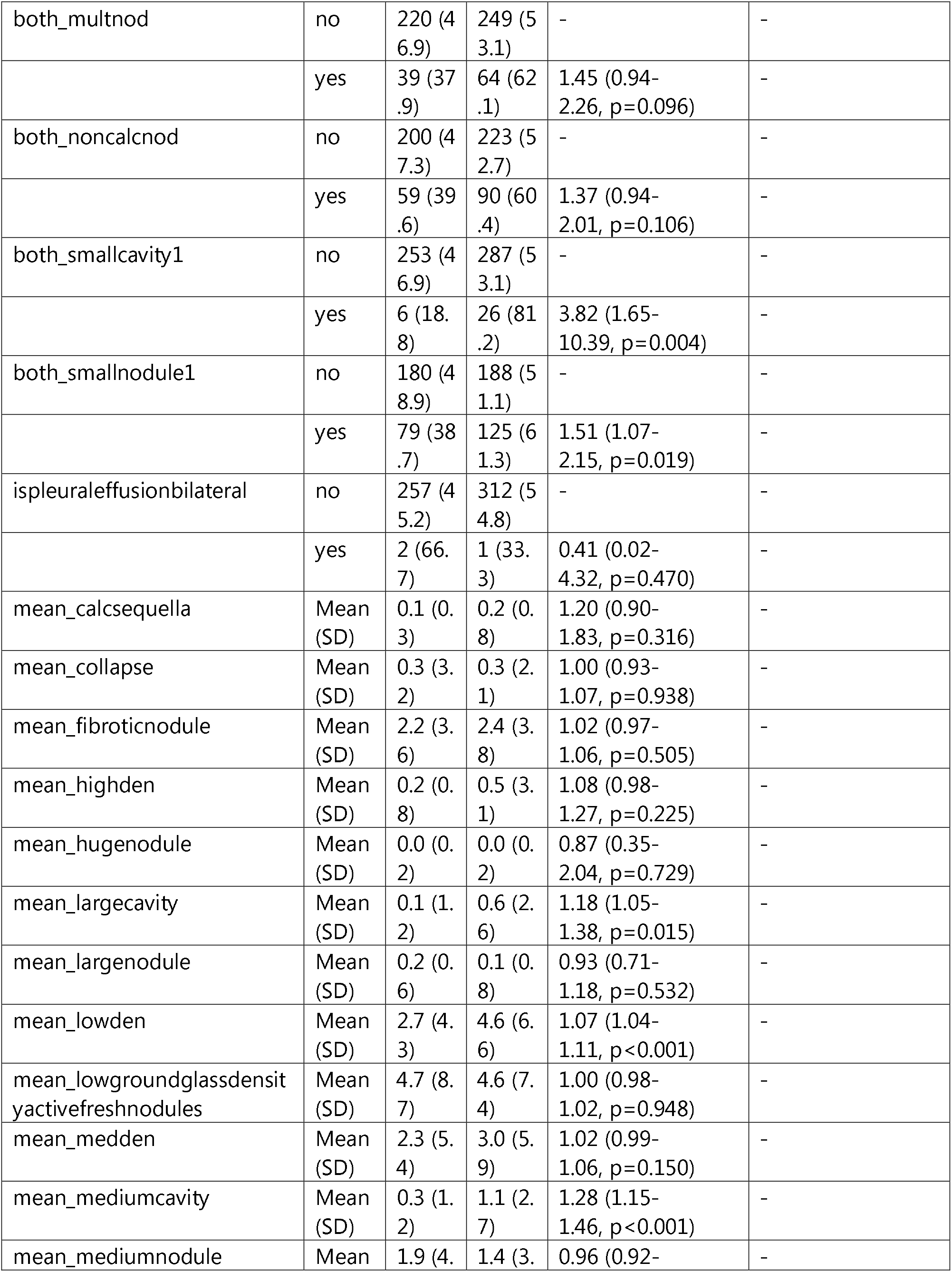
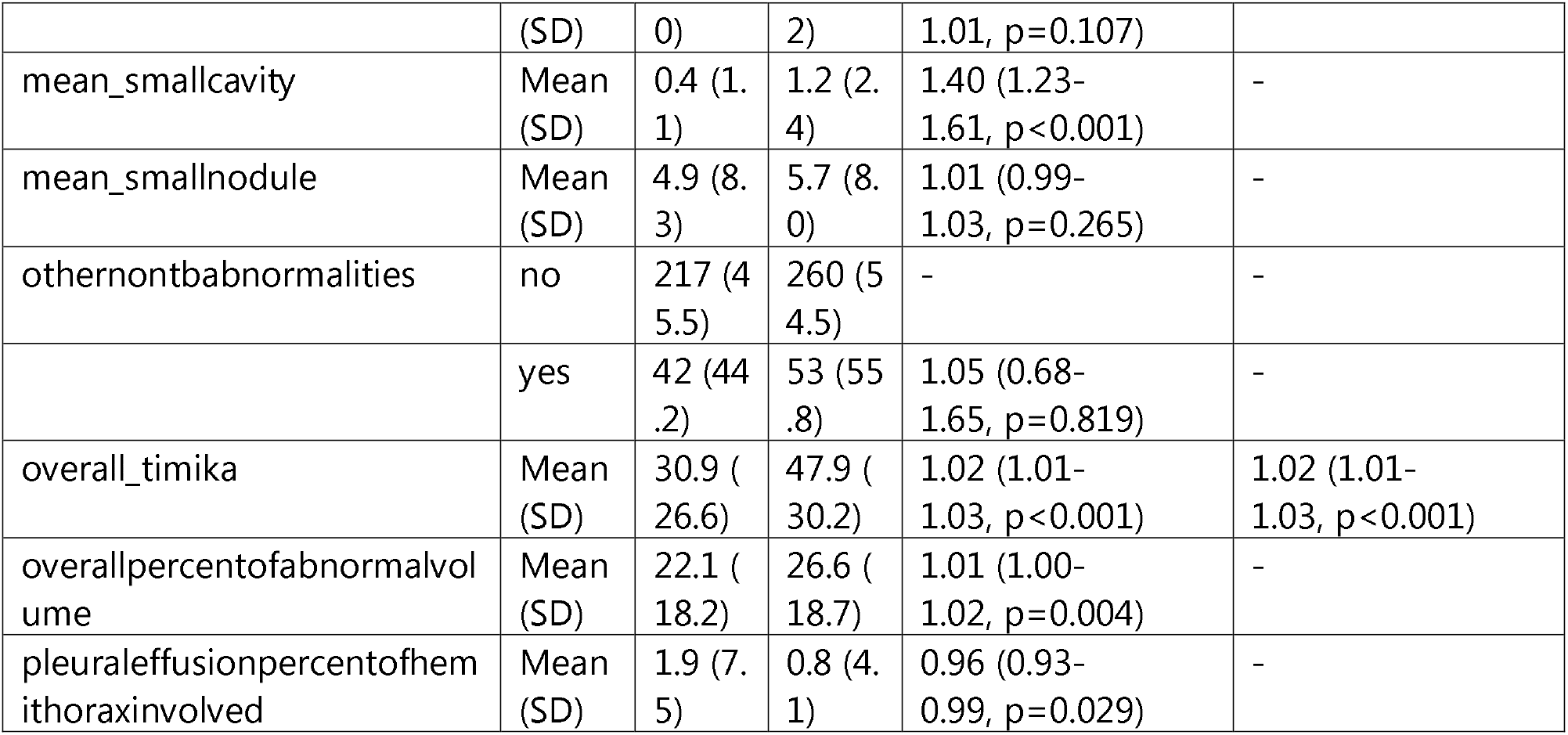
Risk of positive sputum status (1+ or higher) by univariate or multivariate logistic regression analysis on derived features from radiologist observations of CXRs within two weeks of specimens taken prior to treatment. Odds ratios and adjusted odds ratios from univariate and multivariate logistic regression analysis on derived features from radiologist observations of images taken within two weeks of specimens prior to treatment start. Shown are the individual derived features from radiologist observations along with the summary statistics across the prediction categories of Negative or Positive (1 to 9 in 100, 1+, or higher sputum smear microscopy status). Odds ratios with the 95% confidence intervals with unadjusted P-values are shown for each derived feature for the univariate and if applicable, multivariate models. The - sign shows reference categories in the univariate Odds Ratio column or reference as well as excluded variables in the multivariate Odds Ratio column.

### Assessing predictive capacity of machine learning models

The TB portals offers a variety of radiologist observations of CXRs that may provide additional information towards the prediction of baseline sputum smear microscopy status. Therefore, we investigated the additional features comparing predictive performance to Timika Score alone. By doing so, we sought to identify any additional features that might improve upon Timika Score performance and to evaluate how well Timika Score itself could predict sputum status when compared to various feature selection or dimensionality reduction techniques that summarize the radiological features within the data.

#### Comparison of predicting 2+ versus 1+ sputum smear status in training and validation sets

We noted that sputum smear scores of 2+ or greater showed a higher mean Timika Score so we tried two predictive approaches: task one involved predicting positive (1 to 9 in 100, 1+, or greater smear status indicating any active pathogen in the sputum) versus negative smear status while task two involved predicting higher bacterial load positive (2+, 3+, or greater smear status) versus negative sputum status. We split the data into a 70% training set and 30% validation set for the model training and validation. All pipelines were created and run using the MLR3 package which allows for unbiased assessment of model performance by encapsulating the pre-processing steps within the cross-validation approach. All prediction tasks included featureless pipelines showing a non-informative model that only predicts the most prevalent class or randomly selects a class in case of a tie. Thus, the featureless model can be considered a control and predictive models should perform significantly better than this featureless control model.

In the first prediction problem attempting to discriminate positive from negative status, 5-fold cross validation results on the training data showed that most models demonstrated relatively similar predictive performance across pipelines (Table 2). Pipelines using top 5 components by principal component analysis (which captured ∼ 50% of variability in the dataset) tended to show slightly decreased performance in general. Since this prediction problem did not have a large class imbalance, the addition of a class balancing step did not make a significant impact to prediction performance for the pipelines tested. Of note, pipelines only including the Timika Score showed equivalent performance in general to workflows using the top 5 predictive features from various feature selection algorithms. There is a slight decrease in performance of Timika-only models to the best top 5 feature selection model, which reflect that additional features may provide some minimal improvement over Timika Score. To test predictive performance on data which the models had not seen before, we trained the above models on the entire training set and tested on 30% of held out validation data (Table 3). We observed similar findings to the cross-validated results we obtained from the training data.

**Table 2.**
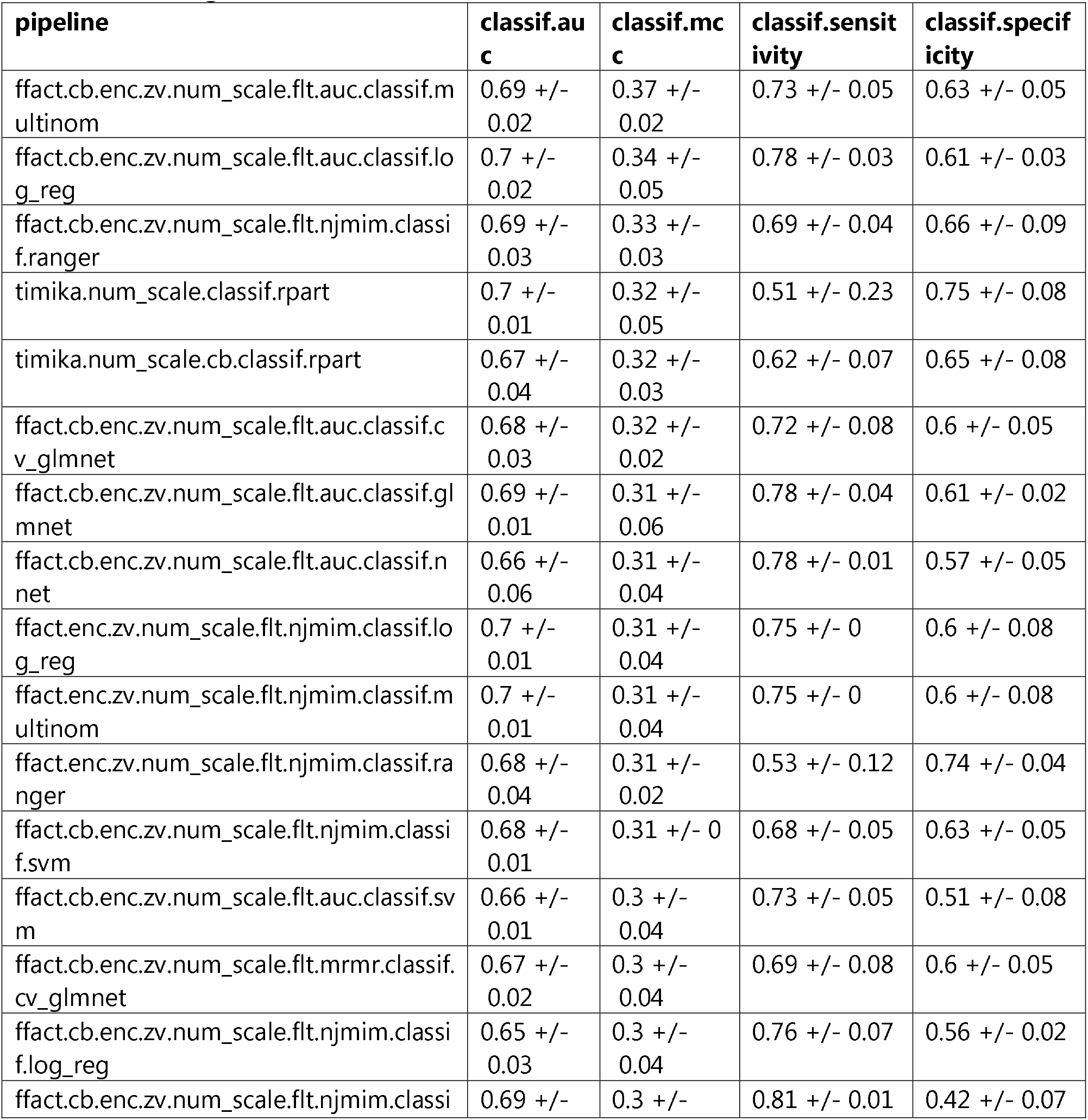
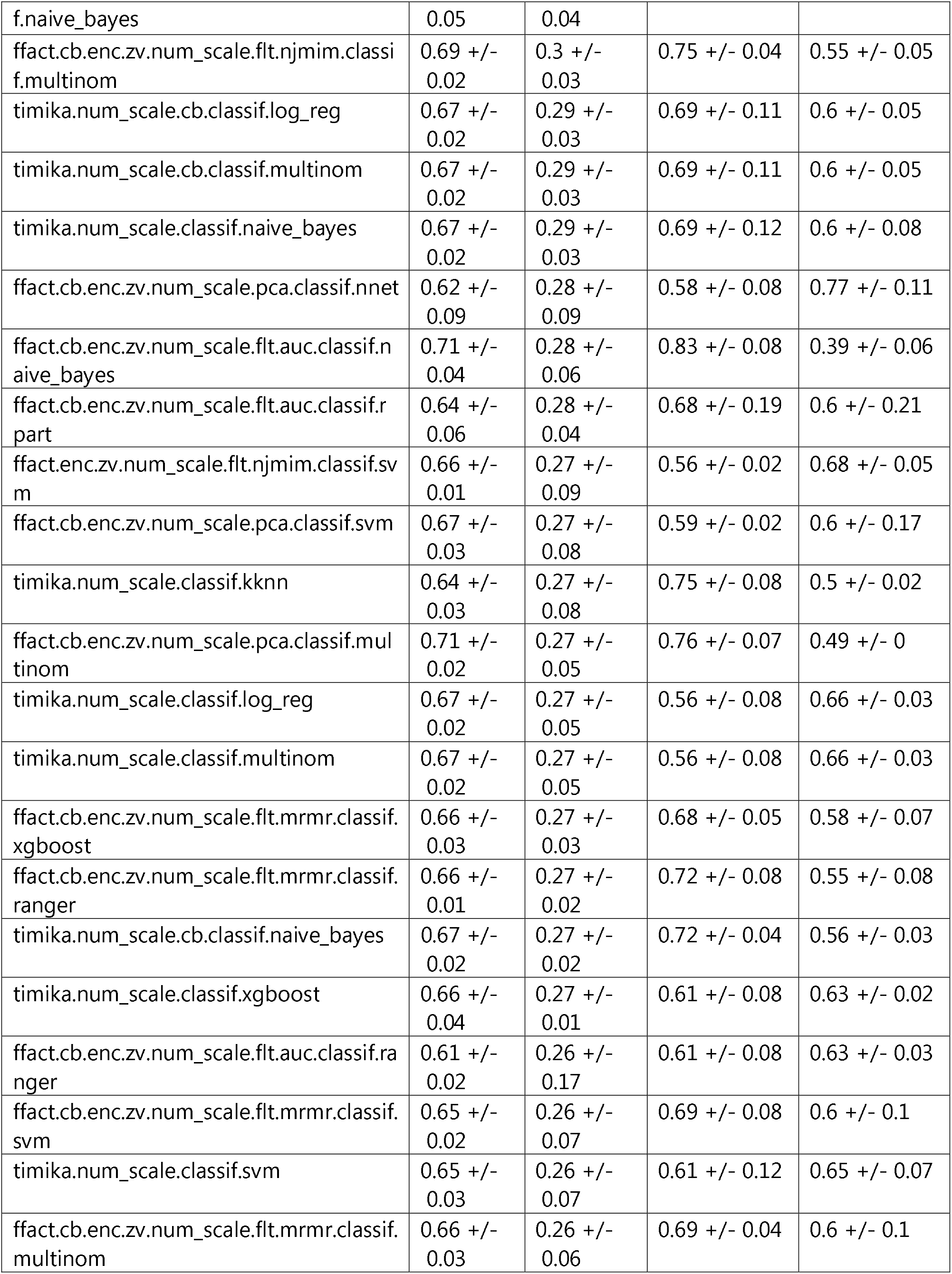
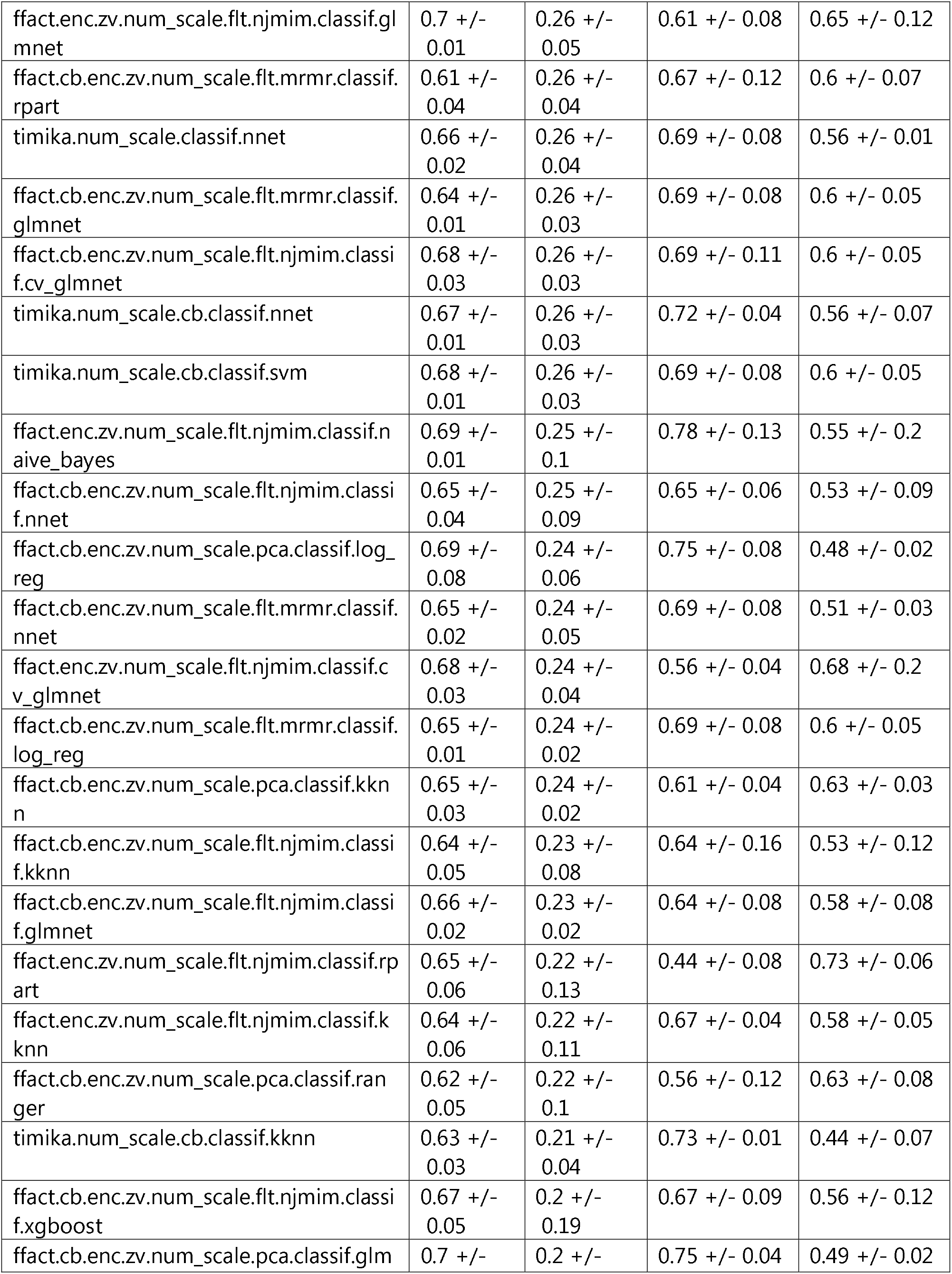
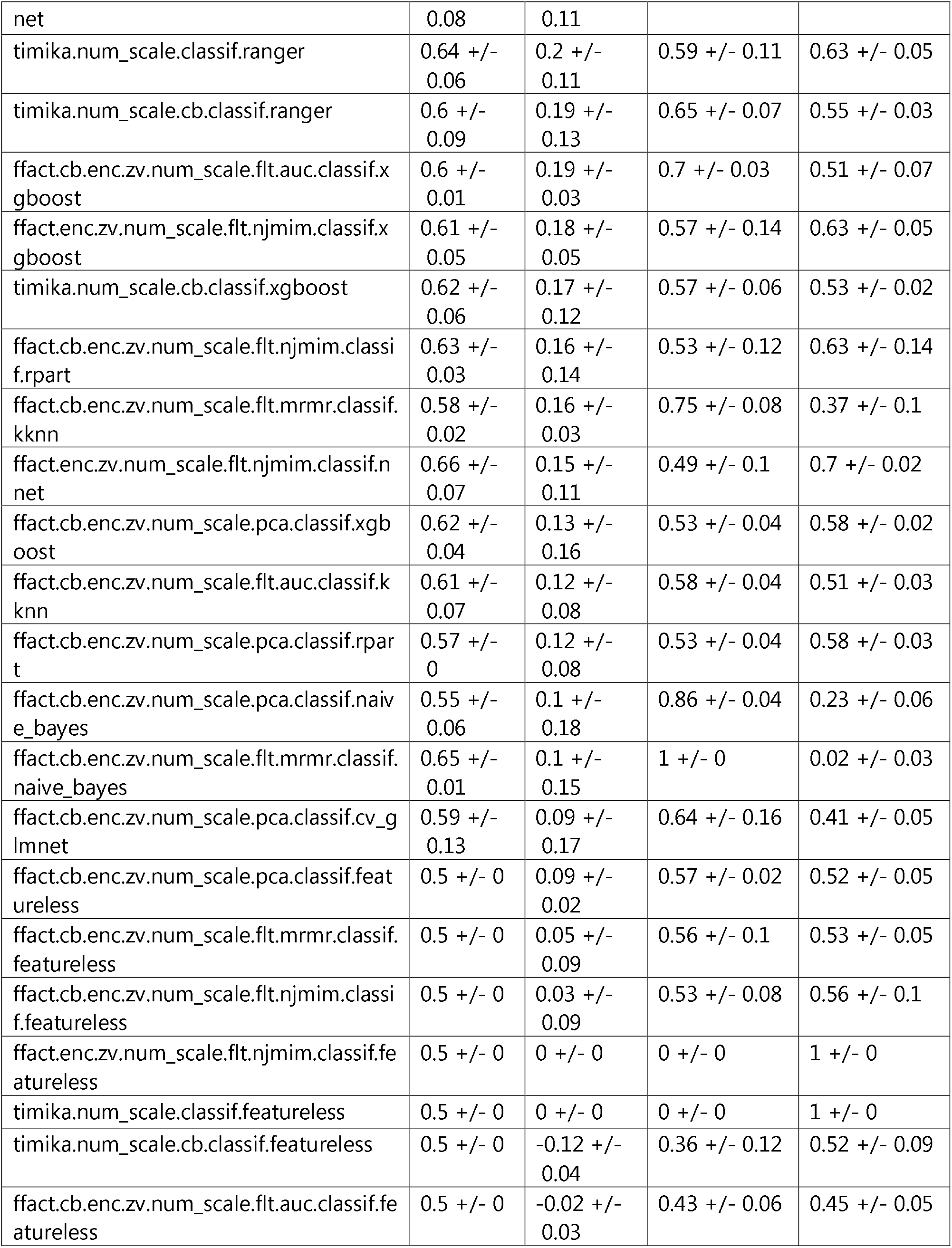
Comparison of machine learning pipeline performance via 5-fold cross-validation for predicting positive (1 to 9 in 100, 1+, or higher) versus negative sputum microscopy status on training data. The machine learning pipeline is shown in the pipeline column. Model performance metrics include Area under the curve (classif.auc), Balanced accuracy (classif.bacc), Matthew’s Correlation Coefficient (classif.mcc), Sensitivity (classif.sensitivity), and Specificity (classif.specificity). Each cell represents the median metric +/- the MAD for the 5-fold cross validation testing on the training data representing 70% of the entire dataset. Pipelines using only the Timika Score for prediction start with Timika in the pipeline name. Each pipeline shows all steps used in the pipeline ending with the machine learning algorithm used and are ordered by classif.mcc.

**Table 3.**
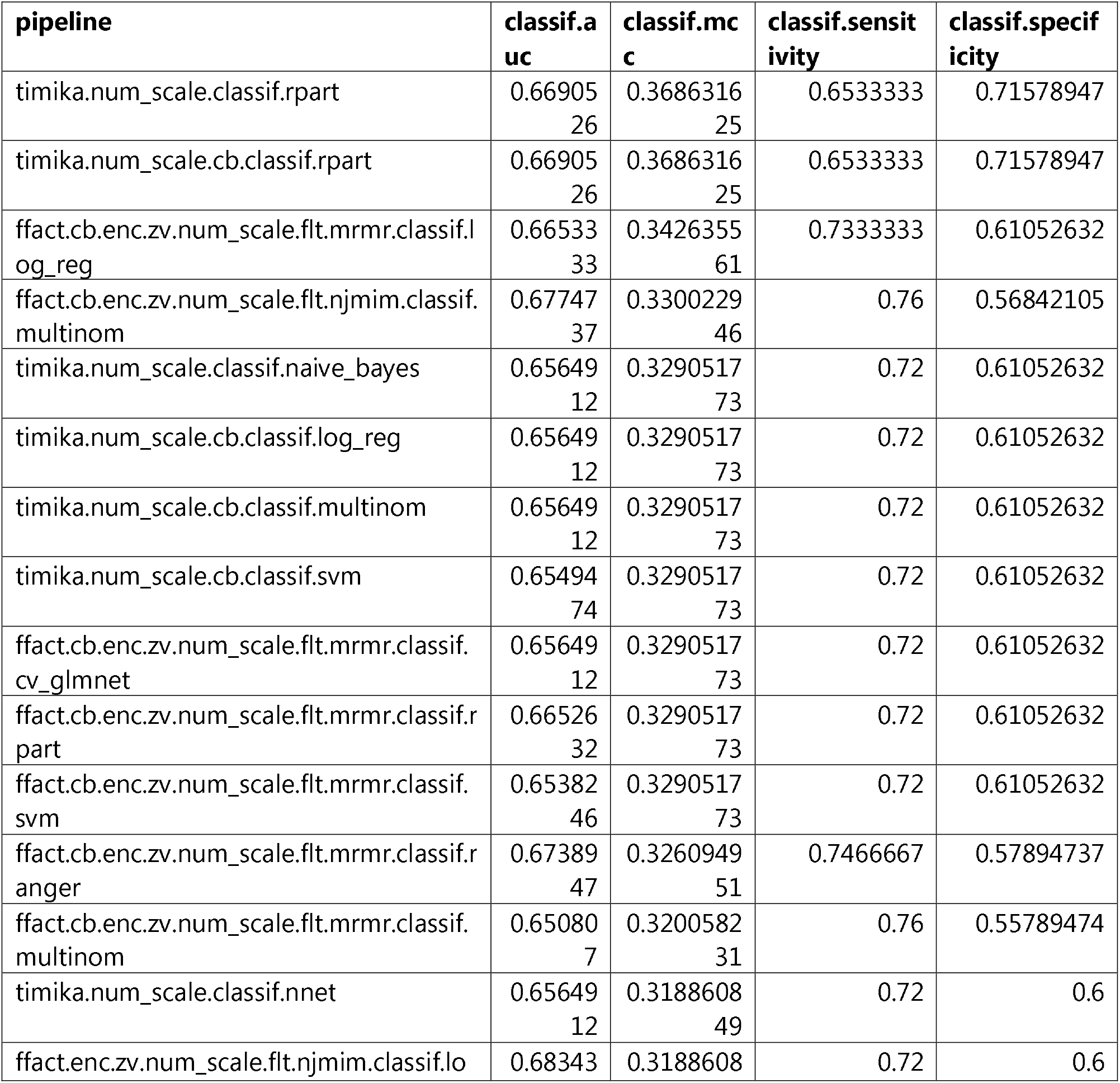
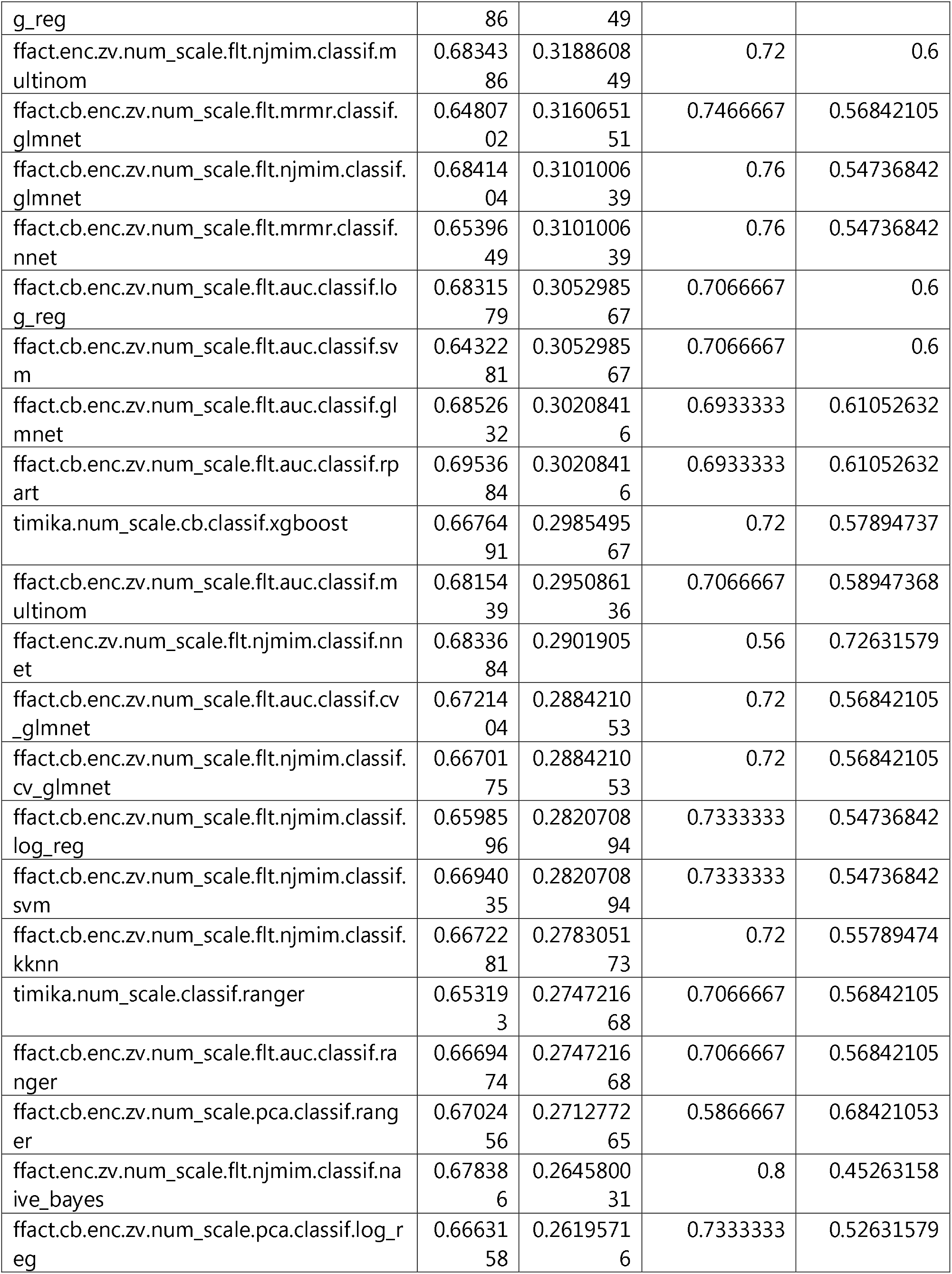
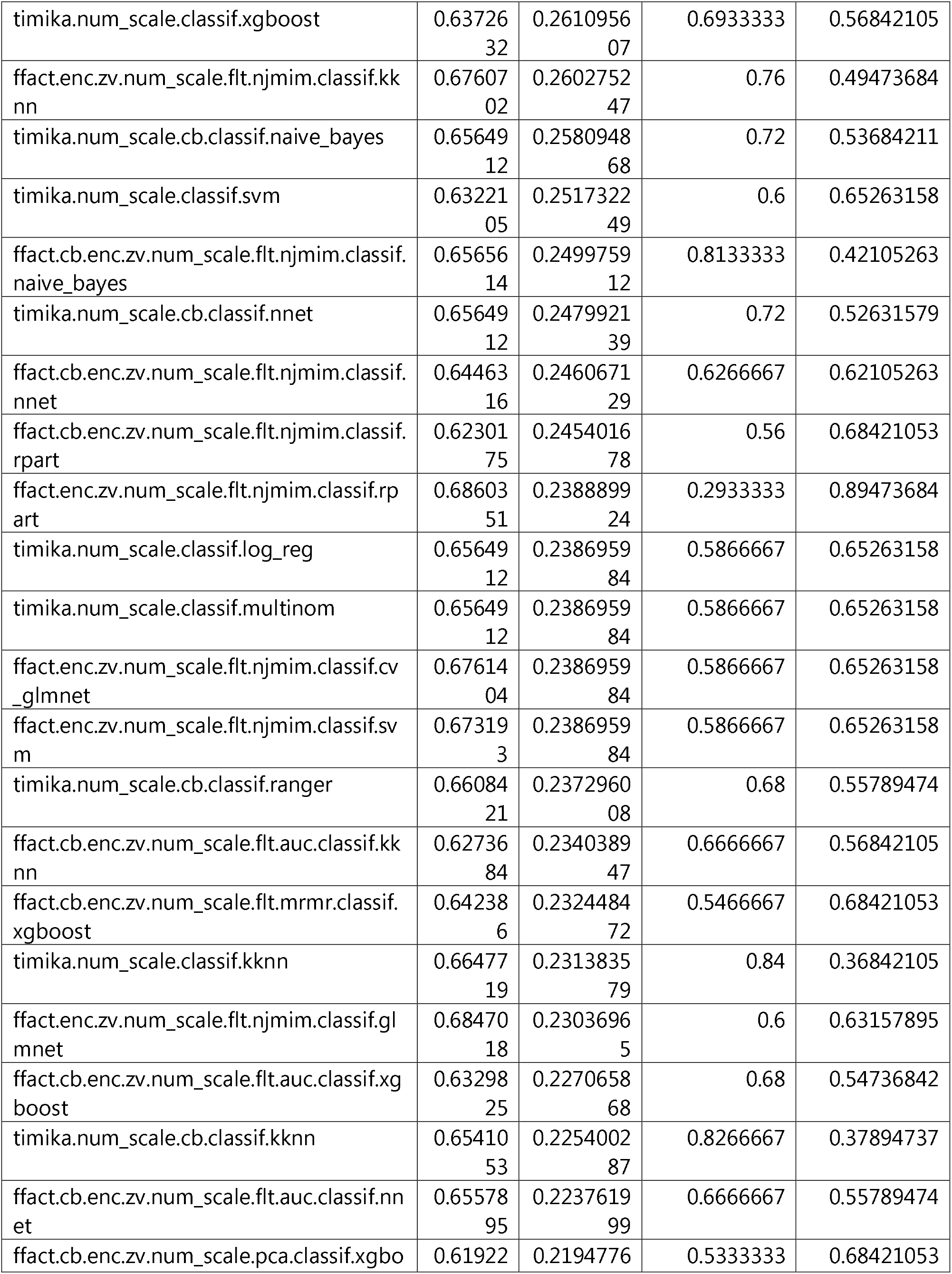
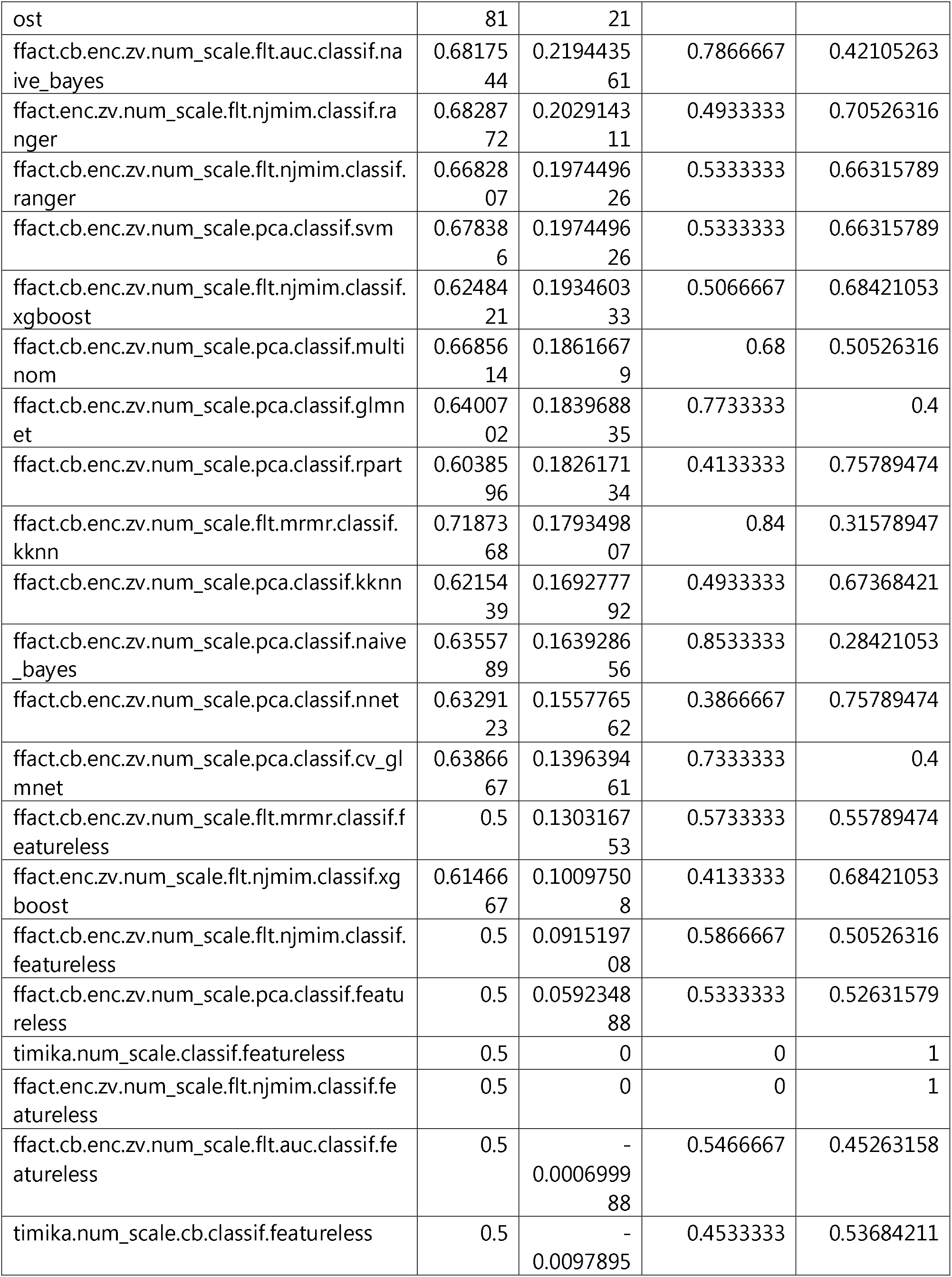
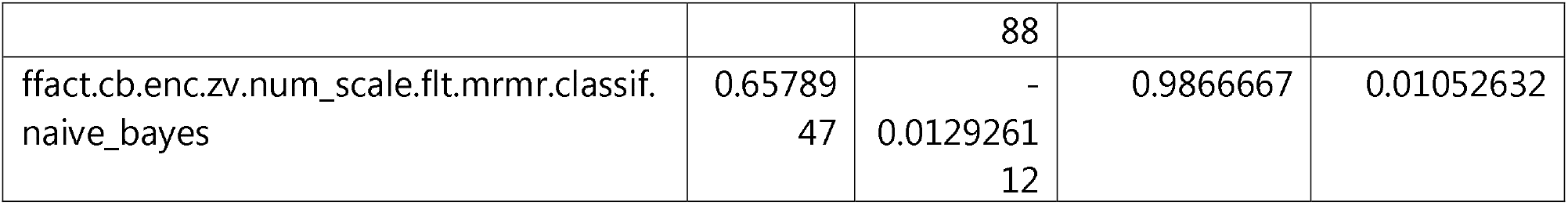
Comparison of machine learning pipeline performance for predicting positive (1 to 9 in 100, 1+, or higher) versus negative sputum microscopy status on validation data. The machine learning pipeline is shown in the pipeline column. Model performance metrics include Area under the curve (classif.auc), Balanced accuracy (classif.bacc), Matthew’s Correlation Coefficient (classif.mcc), Sensitivity (classif.sensitivity), and Specificity (classif.specificity). Each cell represents the metric for the performance on the 30% held-out validation test set after training on the 70% training data. Pipelines using only the Timika Score for prediction start with Timika in the pipeline name. Each pipeline shows all steps used in the pipeline ending with the machine learning algorithm used and are ordered by classif.mcc.

We hypothesized that the second prediction task might demonstrate a performance boost in predictive power since the 2+ sputum status or higher (very high pathogen load in the sputum) showed a larger difference in Timika Score compared to negative. For this prediction task, we removed the borderline 1 to 9 in 100 and 1+ sputum test results from the analysis. 5-fold cross validation results on training set confirmed our hypothesis as we observed increases in performance for reported metrics (Table 4) for both top 5 features pipelines as well as Timika Score only workflows. In general, we observed equivalent performance from Timika Score only pipelines to workflows using the top 5 predictive features from various feature selection algorithms. The benchmarking suggests that while possible to achieve additional gains from the set of derived radiologist observations, these would likely be minimal. Interestingly, though this prediction problem shows a moderate class imbalance, the incorporation of class balancing did not significantly increase or impair performance for the pipelines. When we tested models trained on the entire training set on a validation set of 30% held out data, we saw similar predictive performance to our observation of the 5-fold cross-validated results on the training set (Table 5).

**Table 4.**
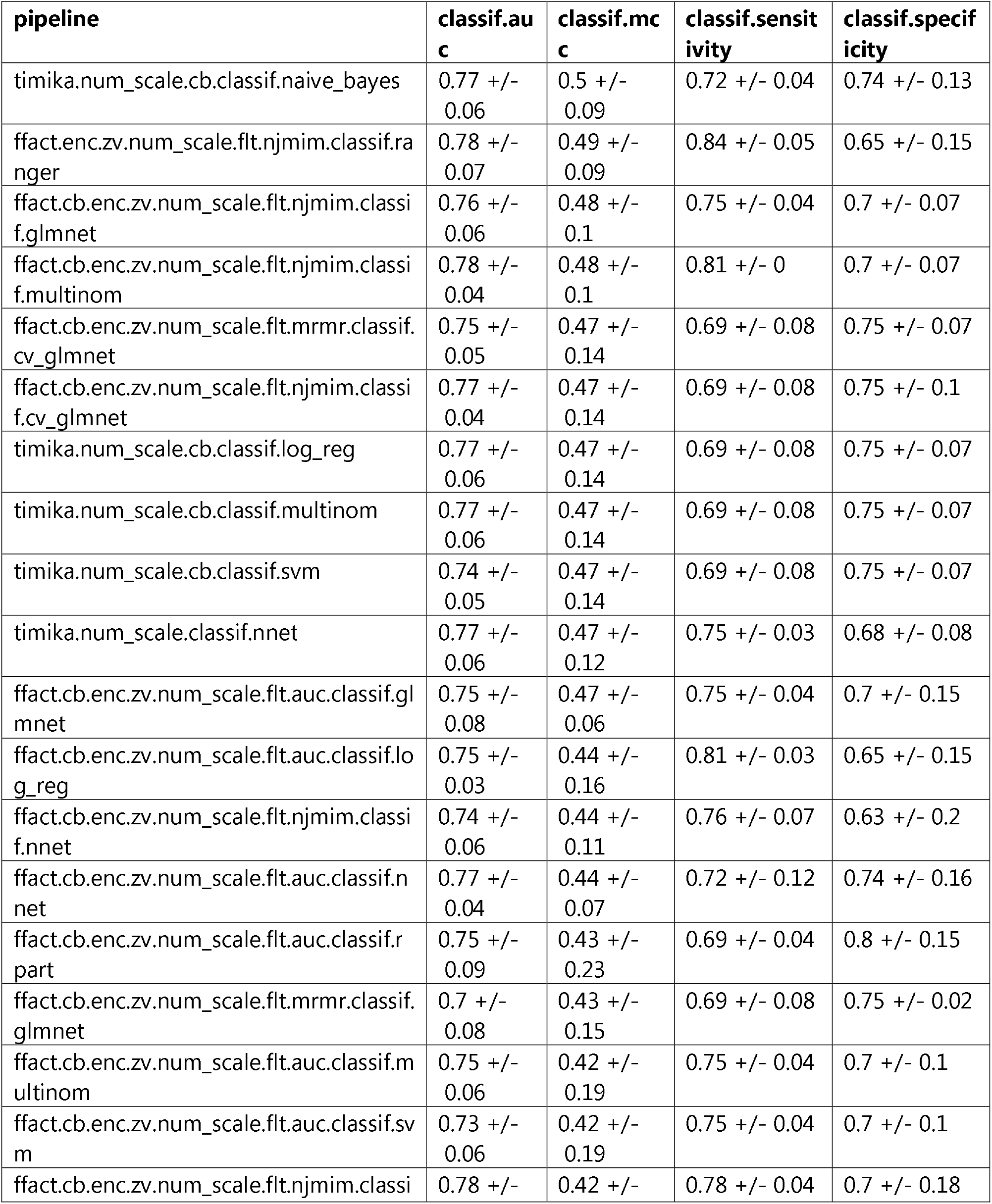
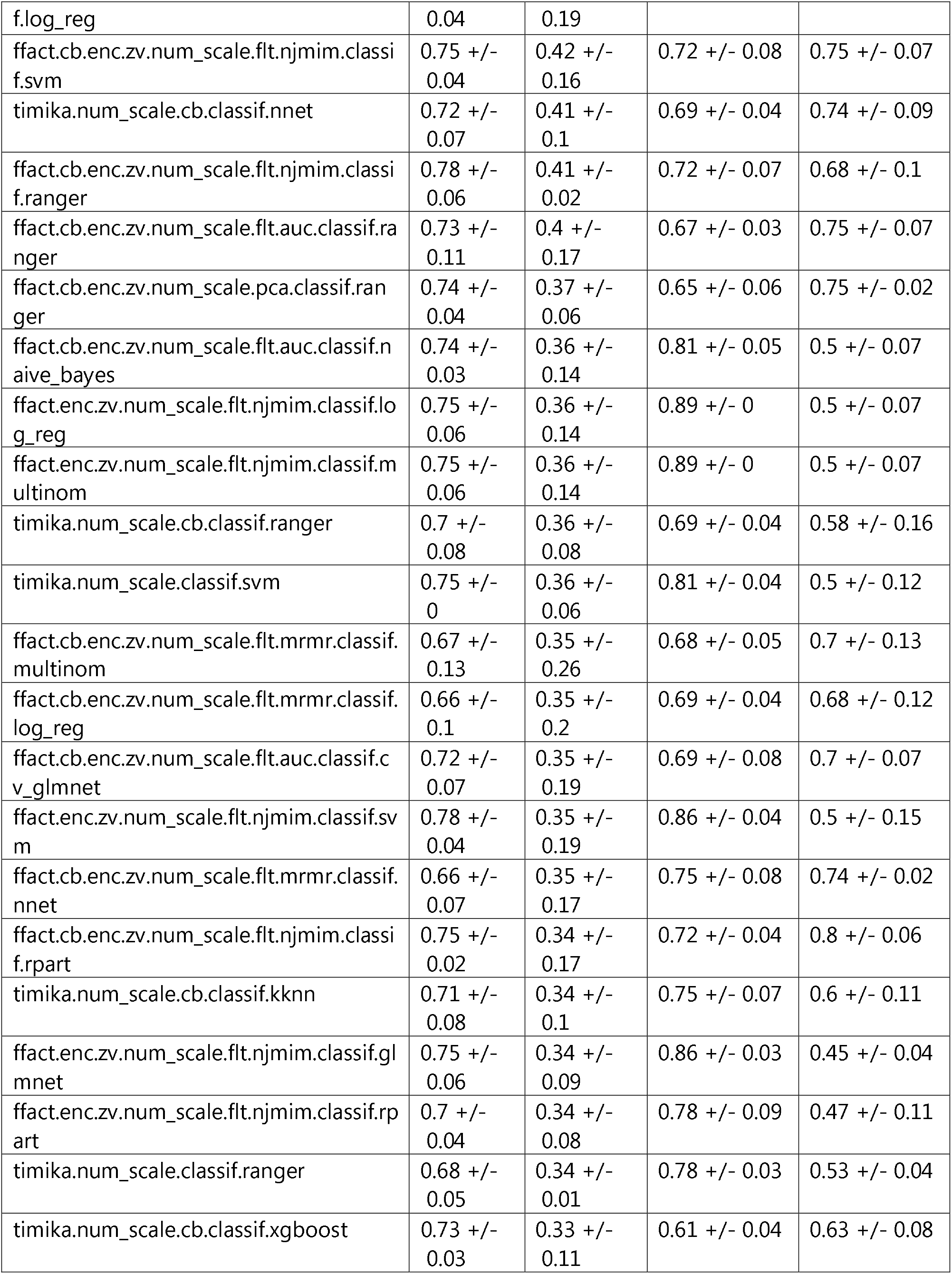
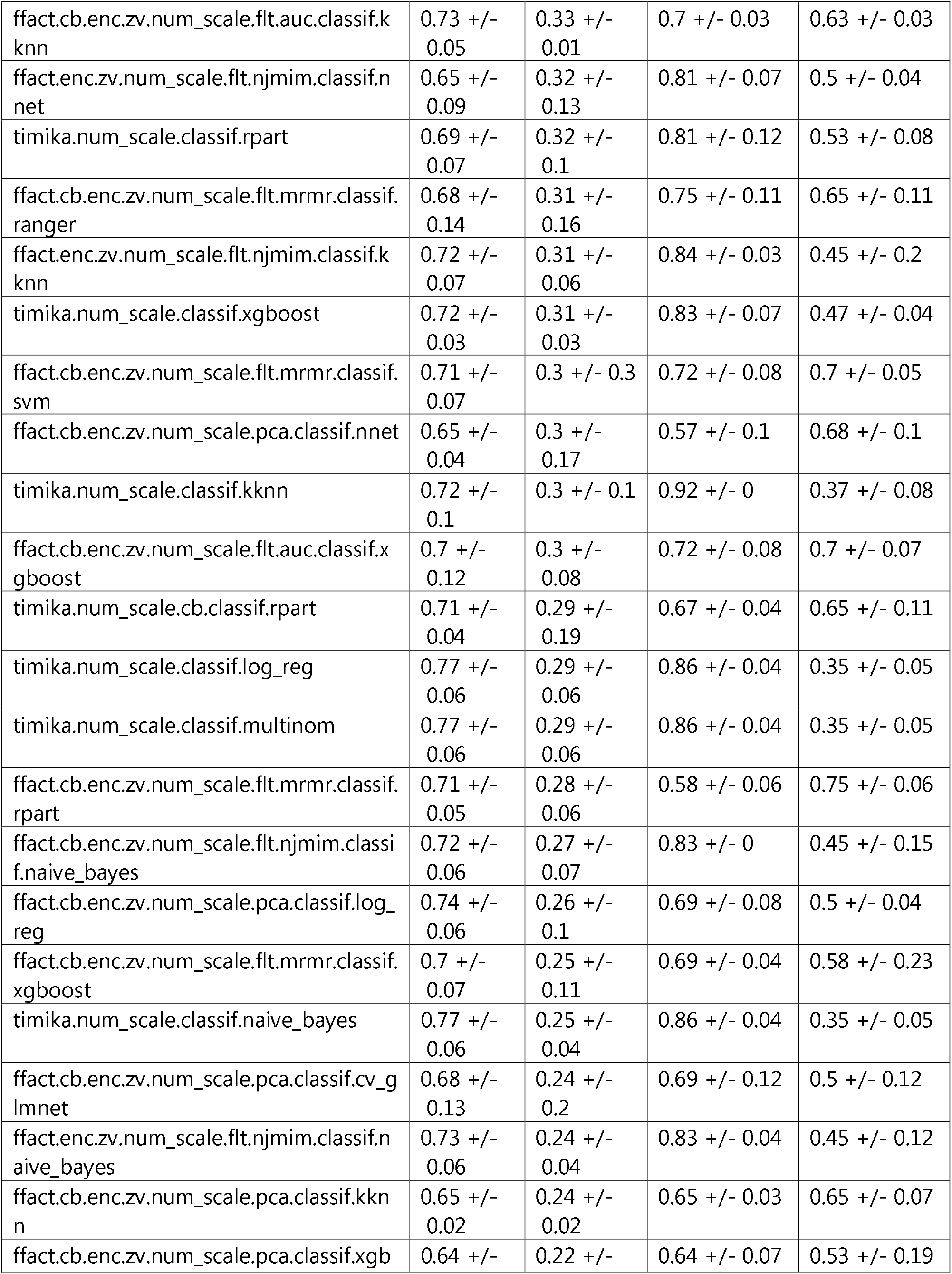
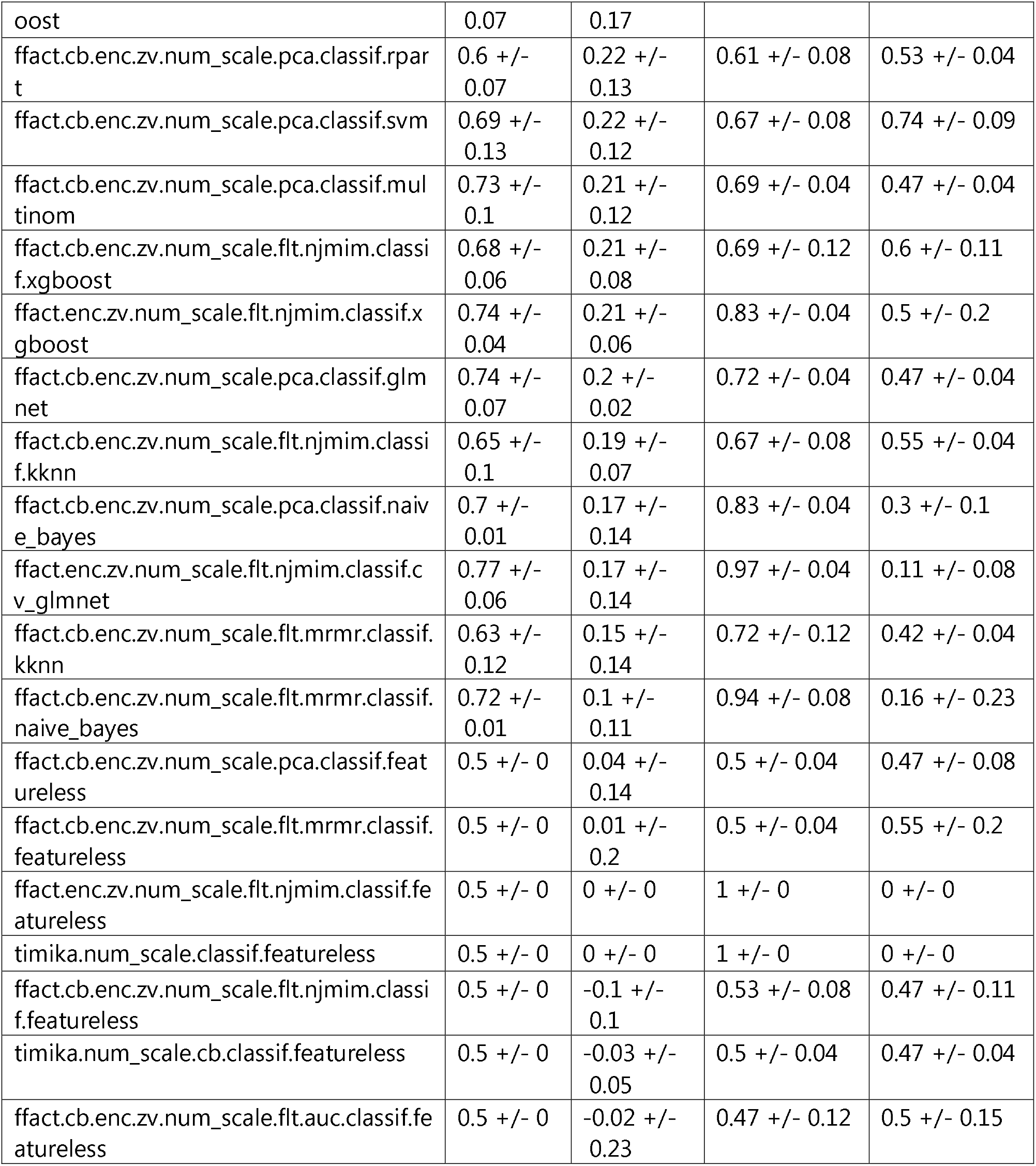
Comparison of machine learning pipeline performance via 5-fold cross-validation for predicting high bacterial load positive (2+ or higher) versus negative sputum microscopy status on training data. The machine learning pipeline is shown in the pipeline column. Model performance metrics include Area under the curve (classif.auc), Balanced accuracy (classif.bacc), Matthew’s Correlation Coefficient (classif.mcc), Sensitivity (classif.sensitivity), and Specificity (classif.specificity). Each cell represents the median metric +/- the MAD for the 5-fold cross validation testing on the training data representing 70% of the entire dataset. Pipelines using only the Timika Score for prediction start with Timika in the pipeline name. Each pipeline shows all steps used in the pipeline ending with the machine learning algorithm used and are ordered by classif.mcc.

**Table 5.**
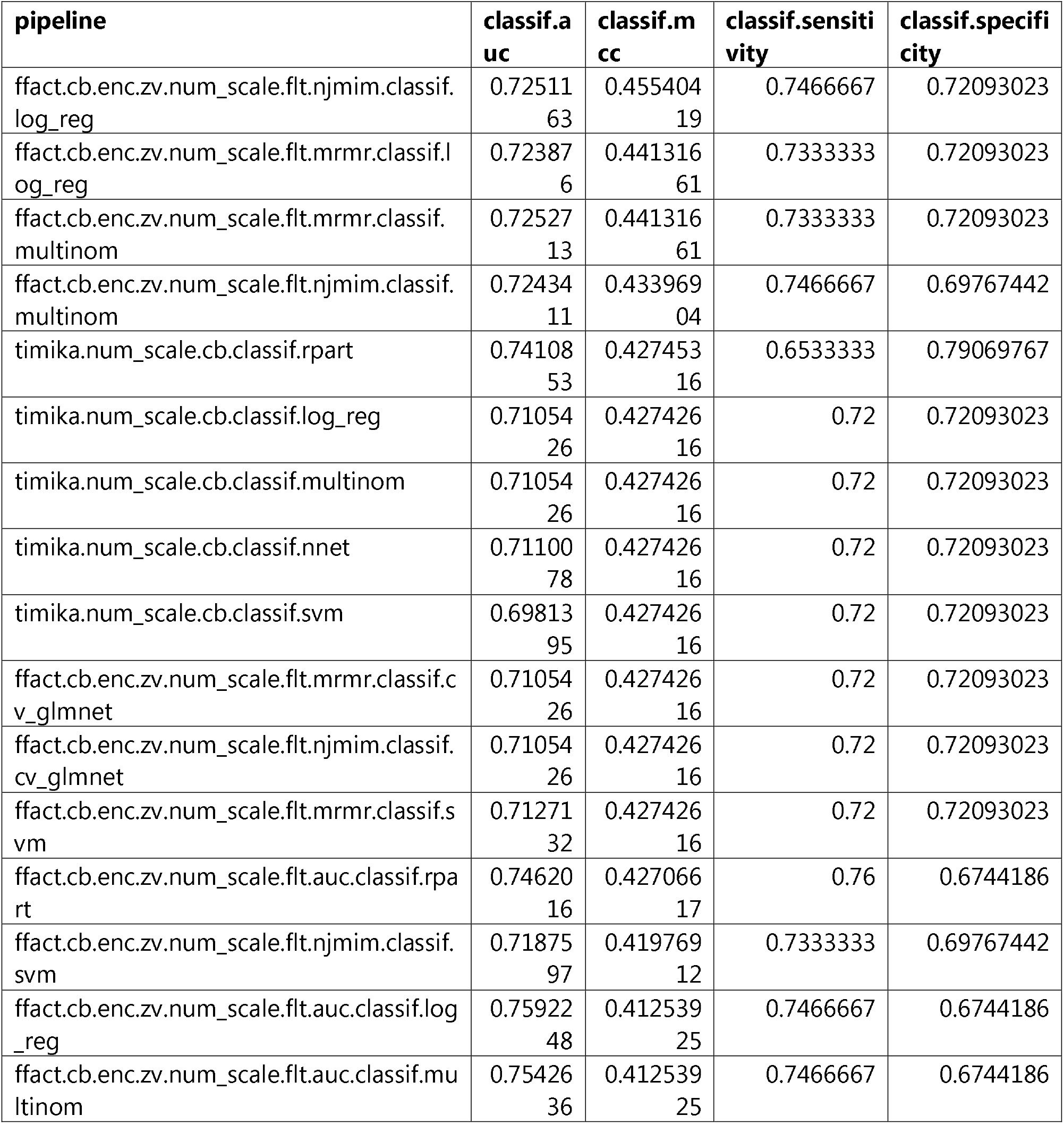
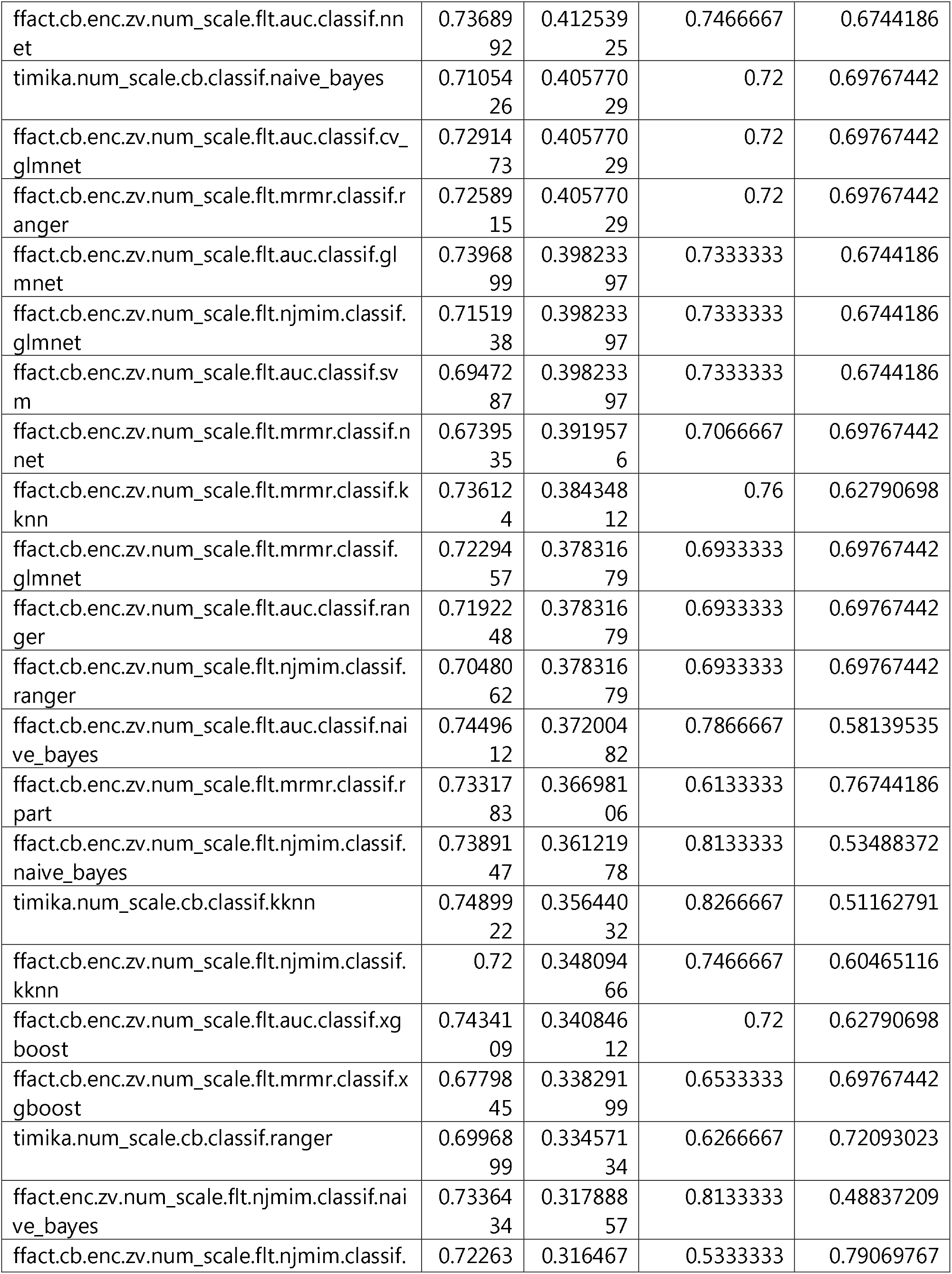
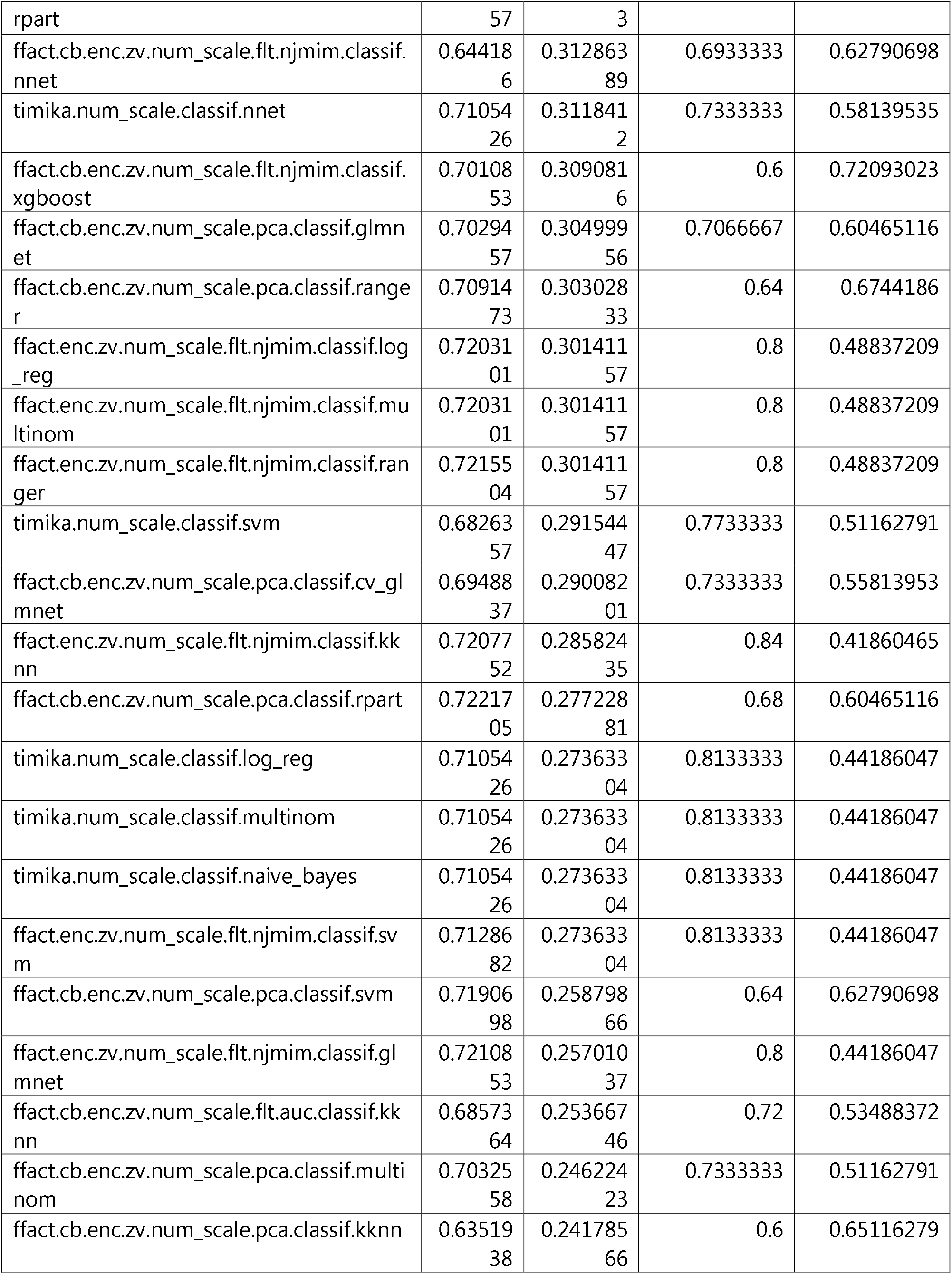
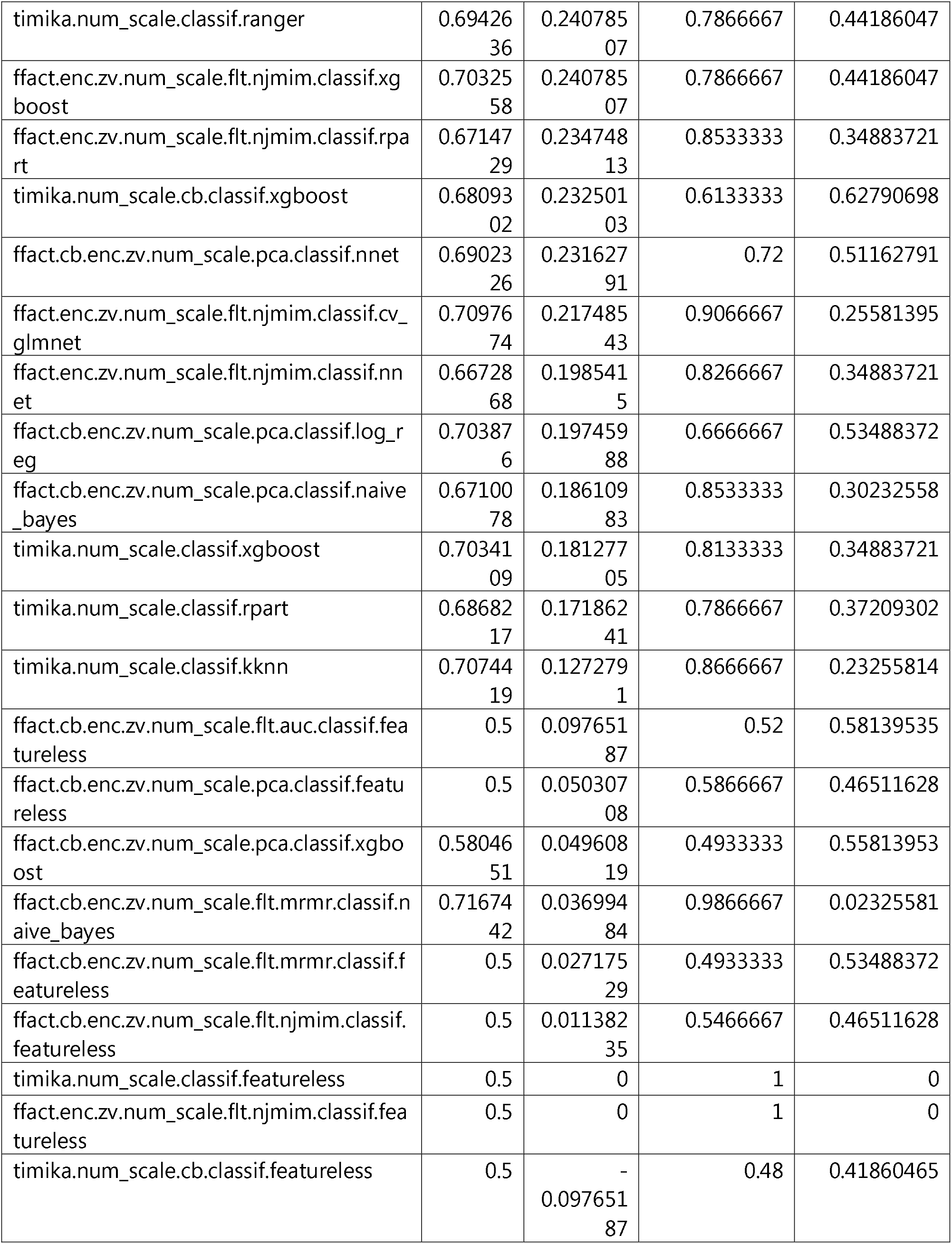
Comparison of machine learning pipeline performance for predicting high bacterial load positive (2+ or higher) versus negative sputum microscopy status on validation data. The machine learning pipeline is shown in the pipeline column. Model performance metrics include Area under the curve (classif.auc), Balanced accuracy (classif.bacc), Matthew’s Correlation Coefficient (classif.mcc), Sensitivity (classif.sensitivity), and Specificity (classif.specificity). Each cell represents the metric for the performance on the 30% held-out validation test set after training on the 70% training data. Pipelines using only the Timika Score for prediction start with Timika in the pipeline name. Each pipeline shows all steps used in the pipeline ending with the machine learning algorithm used and are ordered by classif.mcc.

#### Comparison of best performing feature selection and Timika-only models by bootstrapping

Though our initial training and validation testing suggested that Timika Score only pipelines showed minimal differences with the top 5 feature models, we wanted to further test this for statistical significance. We chose the best performing class-balanced pipelines from the 5-fold cross validated results obtained from the training data. Thus, the best top 5 feature pipeline, Timika Score only pipeline, and a featureless pipeline were selected for further testing. The featureless workflow is a control that would reveal the density of results expected due to random chance regardless of any upstream preprocessing given that classes are balanced. We perform a bootstrapping without replacement (N = 200) on the entire dataset using 70% of the data for training and 30% for testing on each split.

The bootstrapping result on the prediction problem attempting to distinguish positive from negative sputum results showed that the performance of the best top 5 feature pipeline was slightly better than the Timika-only pipeline. Both showed significantly improved performance from the workflow using a featureless model (Fig 4A). The bootstrapping on the prediction problem attempting to distinguish higher bacterial load sputum results (2+ or higher compared to negative) did not show any difference in performance for the best Timika-only pipeline compared to the best top 5 workflow (Fig 4B). As before, both pipelines performed significantly better than the workflow using the featureless model. These results are consistent with the idea that Timika offers generally equivalent predictive performance to using the top 5 features from the dataset. Inclusion of other features may offer minimal improvements in predictive performance depending upon the model or set of features selected.

**Fig 4.**
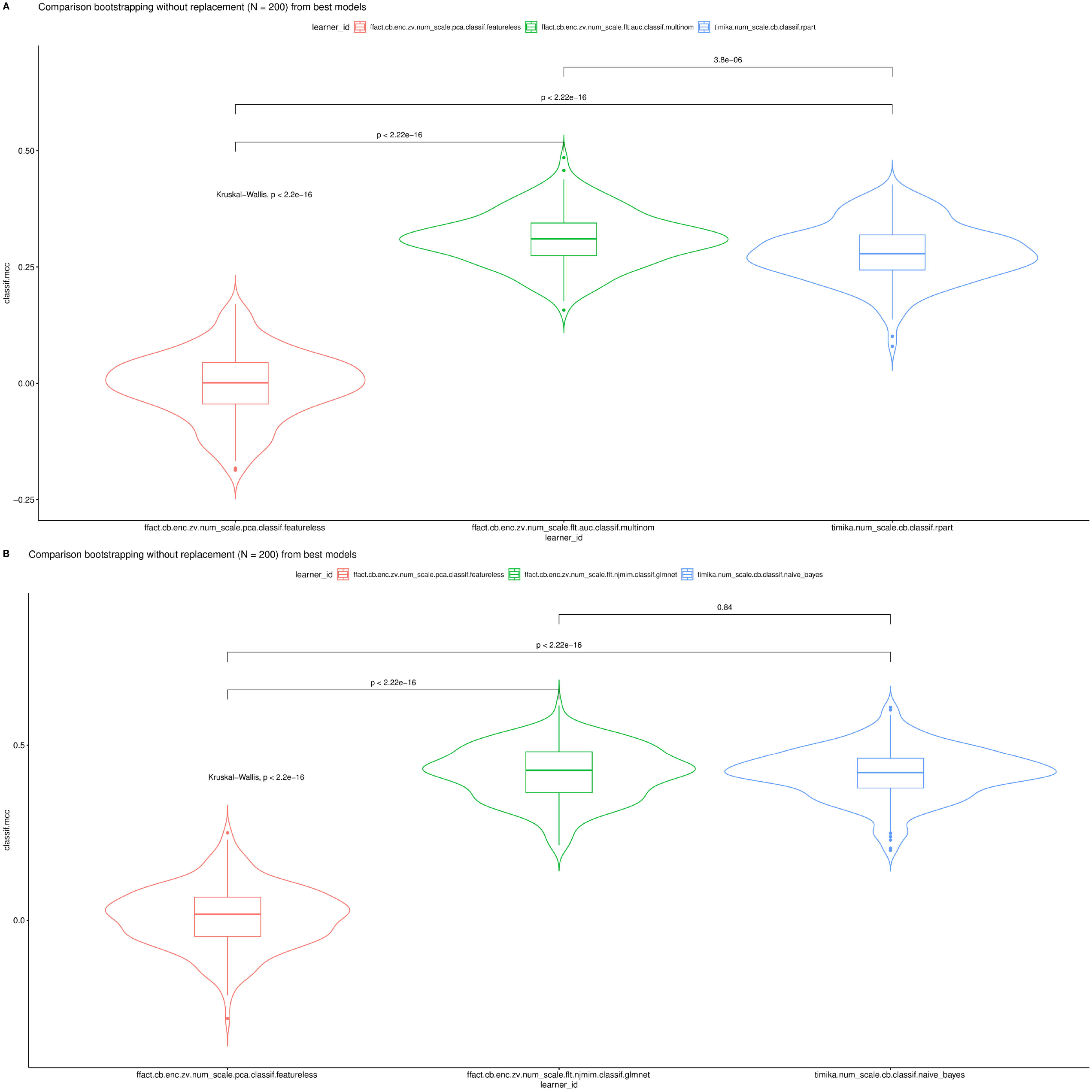
Comparison of best class-balanced pipelines via bootstrapping without replacement (N = 200). The best class-balanced pipelines via 5-fold cross-validation performance on training data were selected for comparison to assess using top 5 features via feature selection or dimensionality reduction as compared to using only Timika Score. A featureless pipeline is used as a control to show expected performance via random selection of outcome. Box plots with interquartile range are overlaid on density plots showing the density of results of Matthew’s Correlation Coefficient across all bootstrapping results per pipeline. Performance is compared across all groups by Kruskal-Wallis and by individual pairs using Wilcoxon test which are shown by the brackets. In A), the performance of the best pipelines for predicting positive (1 to 9 in 100, 1+, or higher) versus negative is shown whereas B) the performance of the best pipelines for predicting high bacterial load positive (2+ or higher) versus negative is shown. Interestingly there is a statistically significant difference between top 5 feature pipeline versus Timika Score pipeline in A) and no significant difference in performance for Timika Score pipeline in B). This shows that additional derived features from radiologist observations may result in small performance gains although Timika Score alone provides generally equivalent prediction performance for the identified best models.

## Discussion

X ray imaging is a useful approach to diagnose and monitor disease progression and status during routine TB clinical management. X ray imaging cannot discern the type of resistance of tuberculosis as well as characterize the amount of pathogen in sputum, which only microbiological methods can provide. These approaches assist clinicians with a more complete understanding of the case and understanding their relationship is important. CXR is relatively less expensive than other imaging modalities such as CT permitting its wider use especially in LMIC that may face challenges with infrastructure cost to support routine CT use. Using CXRs, radiologists can report on a variety of observations that determine lung biomarker status such as overall abnormal volume of the lungs, presence of cavity, and presence of nodule, which the TB Portals resource collects, standardizes, and provides as part of the patient record. Here we investigated the previously reported Timika Score that can be derived from CXR radiologist observations to characterize pre-treatment severity of disease. TB portals provides a unique real-world repository of TB cases, especially drug resistant cases to bridge across distinct domains including radiological, pathogen genomic, microbiological, and clinical features; it is especially suited to serve as a large reference resource for assessing derived scores like Timika Score for testing in a real-world database of especially challenging TB cases. Our goal was to assess the plausibility and utility of the derived Timika Score within this real-world resource by studying its relationships to the other available case characteristics.

We demonstrate that Timika Score associates with other case characteristics consistent with prior reporting of TB clinical risk factors. For instance, we show that images from patients with a lower BMI tended to have a higher Timika Score and less of a change from the initial CXR to the last available CXR. TB and BMI have been reported to show a strongly logarithmic association and there was reported fivefold increase in age-adjusted incidence of new pulmonary TB in lowest BMI group compared to highest in a study of 1.7 million Norwegians (18, 19). In the same report, Tverdal mention an interesting U-shaped association with BMI and all-cause mortality which is strikingly like the U-shaped association we observed with Timika Score and BMI. In our analysis, increasing age tended to associate with higher Timika Score and less of a change from the initial CXR to the last available CXR. This is consistent with higher mortality, morbidity, and risk of TB with increasing age especially since the symptoms of TB may be confused with other age-related illnesses (20) resulting in delayed diagnosis or treatment. Moreover, when comparing Timika Score with other clinical factors associated with the case, we observe both higher Timika Scores and lower relative changes in Timika Score in cases with higher-risk clinical factors (XDR, Relapse, etc.) or poor reported outcomes (e.g. Treatment failure, Died, etc.). This is consistent with prior reports examining predictors that affect change in radiological lesions over the course of treatment monitoring (13).

Finally, we demonstrated that Timika Score show a clear and statistically significant predictive capacity for baseline, pre-treatment sputum microscopy status in the cohort of new cases we identified from the TB Portals repository. This is important because the original reports on Timika Score suggested the same association on a smaller dataset (15) that did not span across the wide-range of participating sites from 14 countries. Taken together, these observations support that the Timika Scores we are calculating reflect lung biomarker status consistent with accumulated knowledge of the radiological and clinical associations in TB disease.

This analysis has several limitations and caveats when interpreting results. The TB Portals is a real-world data repository to better understand DR TB so it is challenging to separate identified associations with other observed or unobserved variables from the case. Moreover, respective images and test results for each case are not collected uniformly in time but rather as clinical management of the case allows. We select cases for inclusion into the analysis cohort based upon criteria we believe will accurately represent associations between images and microbiological test results but we cannot rule out timing or other aspects of the case impacting the associations we observe. For example, we noted both lungs involvement of calcified nodule as showing higher risk of pre-treatment sputum positivity. Such a marker suggests a long-term prior history of pulmonary TB that might not be reflected in the “New” case definition from WHO. Collecting a prior history of chronic lung symptoms around the baseline sample collectiong might allow us to see if the relationships we identified remain after stratifying by these symptoms that could suggest a period of prior disease burden. Given these caveats, the modeling and visualizations need to be interpreted as hypothesis-generating.

A key goal of this study was to identify a risk score (either new or previously reported) that could encapsulate a temporal snapshot of case dynamics relating to disease risk such as poor outcome or infectiousness. The CXR derived Timika Score may provide a useful score in this regard from the initial testing we performed. One caution with regards to the utility of Timika Score from this analysis is that the risks and associations with sputum microscopy positivity were calculated for samples taken prior to treatment. The dynamics of microbiological status in sputum might not reflect the same dynamics of lung biomarkers in response to treatment. Given these dynamics, applying the same risks from Timika Score for sputum positivity (i.e. presence of pathogen in sputum) after treatment is not advisable and may require new approaches that stratify by type of resistance and different treatment regimens. It also may require additional data collection to support the requisite number of cases given the real-world nature of the resource where only subsets of cases may meet the inclusion and exclusion criteria for analysis.

The temporal relationships between lung status (as observed in CXR images) and bacterial pathogen load in the sputum (as observed by microscopy) are complex. For instance, at the beginning of disease, radiological features may not be detectable in the lungs despite the presence of bacteria in sputum. Meanwhile, towards the end of treatment, sputum may no longer contain TB pathogen indicating a non-infectious case; however, the pathogen may remain in certain areas of the lung such as cavities. Given these intricacies, additional research is warranted to determine if improvements can be made in Timika Score to account for these situations. Moreover, there may be limitations to the granularity of a clinical score like Timika that can be generated from CXRs given the limitations of the modality with regards to imaging detail. It may be necessary for other modalities such as CT to account for more complex features such as the size of cavities, nodules or other aspects identified in the lung, which may not be obvious on a CXR. The assessment of clinical scores such as Timika Score coming from these various modalities is especially important for hard-to-treat cases such as MDR and XDR TB where scores can be compared in the context of other features of the case.

We observed the best predictive performance in models predicting higher bacterial load sputum status. This improvement in predictive performance for high bacterial load sputum statuses (e.g. 2+ or higher) illustrates the nuances associated with predicting sputum status from Timika Score. The borderline cases such as 1 to 9 in 100 or 1+ are more challenging to predict as pathogen load is only slightly higher than the negative status specimens and furthermore some negative samples may be false negatives. These false negatives may suffer from issues such as sensitivity due to the challenges of acquiring a usable sputum sample. Despite these nuances, our results confirm prior reported Timika Score utility for predicting baseline sputum positivity albeit with better performance for high-bacterial load sputum samples. The high pathogen load cases would also be among the most infectious and challenging to treat; therefore, we were satisfied to observe the higher predictive performance in this clinically important group. We believe that adding CXR-derived Timika Score to the TB Portals resource will open opportunities to other researchers to utilize this score to understand TB in a real-world setting. The score can serve as a reference from which to test against additional clinical scores that could be derived from the available set of features captured in TB portals.

## Supporting information

STROBE checklist

## Data Availability

The TB portals program necessitates all users of the data sign a DUA before access to the underlying, de-identified clinical data is provided and the data can be requested at the following URL (https://tbportals.niaid.nih.gov/download-data). Therefore, this study provides the code to reproduce the analysis without the underlying raw data (https://github.com/niaid/tbportals.xray.sputum.2021) in compliance with the DUA. To aid reproducibility, the list of public identifiers of the cases is provided in Supplementary Table 4.

## Acknowledgements

We would like to thank Qinlu Wang, Jingwen Gu, and Ziv Yaniv for helpful suggestions; the MLR3 team for development of the MLR3 suite of packages. This research was supported in part by the Office of Science Management and Operations (OSMO) of the NIAID. For their contributions to the vision and requirements of TB Portals, we would like to thank: Mike Tartakovsky, Darrell Hurt, Alina Grinev, and members of the TB Portals team.

## Supporting Information

**S1 Table. Case characteristics of the cohort of cases selected for evaluation of baseline sputum microscopy status (N = 572).** Case characteristics were compared by baseline sputum microscopy status. P-values were calculated for continuous variables (age_of_onset, bmi, overall_timika) using analysis of variance test. P-values for categorical variables (registration_date, gender, country, type_of_resistance, outcome, current_smoker) were calculated using Chi-squared test.

**S2 Table. Case characteristics of the cases with CXR radiologist observations used for assessing Timika Score in relation to other case characteristics (N = 1761).** Cases in the TB portals publicly shared dataset having a CXR with available radiologist were selected. The case characteristics are shown.

**S3 Table. CXR derived features from radiologist observations in the cohort of cases selected for evaluation of baseline sputum microscopy status (N = 572).** The derived features from the available radiologist observations from the cohort of selected cases used for evaluation of baseline sputum microscopy status were compared by baseline sputum status. P-values were calculated for continuous variables (mean_collapse, mean_smallcavity, mean_mediumcavity, mean_largecavity, mean_lowden, mean_medden, mean_highden, mean_smallnodule, mean_mediumnodule, mean_largenodule, mean_hugenodule, mean_lowgroundglassdensityactivefreshnodules, mean_fibroticnodule, mean_calcsequella, overall_timika) using analysis of variance test. P-values for categorical variables (both_lungs, both_collapse1, both_smallcavity1, both_mediumcavity1, both_largecavity1, both_isanylargecavitymult, both_multiplecavitiesbeseen, both_lowden1, both_medden1, both_highden1, both_smallnodule1, both_mediumnodule1, both_largenodule1, both_hugenodule1, both_calcnod, both_noncalcnod, both_clustnod, both_multnod, both_lowgroundglassdensityactivefreshnodules1, both_fibroticnodule1, both_calcsequella1) were calculated using Chi-squared test.

**S4 Table. Patient and condition ids for the cohort of cases selected for evaluation of baseline sputum microscopy status (N = 572).** A table of patient and condition ids is provided for the de-identified records that were used for evaluation of baseline sputum microscopy status.

**S5 Table. Patient, condition, and imaging ids for the cases having CXRs with radiologist observations used for assessing Timika Score in relation to other case characteristics (N = 1761).** A table of patient, condition, and imaging ids with associated relative date of imaging is provided for the de-identified records that were used for Timika visualizations. For images used for temporal analysis of changes in Timika Score over time, the temporal_analysis column provides a filter equal to “yes” to select only those sets of images.

